# SleepJEPA: Learning the latent world of sleep with at-home sleep data to estimate disease risk

**DOI:** 10.64898/2026.03.20.26348834

**Authors:** Benjamin Fox, Joy Jiang, Dung T. Hoang, Elizabeth Brush, Pavel Boulgakov, Sajila Wickramaratne, Ankit Sakhuja, Oren Cohen, Mayte Suarez-Farinas, Neomi A Shah, Ankit Parekh, Girish N Nadkarni

**Author notes:** These authors contributed equally (Ankit Parekh, Girish N Nadkarni).

## Abstract

Sleep disturbances lead to cardiovascular (CV), metabolic, and neurological diseases. While in-lab polysomnography (PSG) is the gold standard for measuring sleep disturbances, at-home PSG (hPSG) are more cost-effective, less resource intensive, and have been extensively used in large-scale studies. Further, hPSG devices that record EEG, EOG, and EMG are developing rapidly and collect similar data compared to in-lab PSG . However, the link between hPSG measurements and future disease risk is not well understood. We present SleepJEPA, a foundational sleep study representation model trained via a joint embedding predictive architecture that learns full night, multichannel sleep representations using hPSGs in the latent space, uncovering high-dimensional information that more precisely informs future health outcomes than standard clinical scoring. SleepJEPA was trained, validated, and tested with 422,035 hours of sleep signa.0l data from 55,518 sleep studies. It accurately estimates 1-to 15-year cumulative risk using a discrete hazard loss function for 10 conditions, including angina (integrated area under the receiver operating characteristic curve at 15 years [*iAUC*_15_] = 0.73), CV disease death (*iAUC*_15_ = 0.83), congestive heart failure (*iAUC*_15_ = 0.85), coronary heart disease death (*iAUC*_15_ = 0.85), incident cognitive decline (*iAUC*_10_ = 0.65), diabetes (*iAUC*_10_ = 0.82), hypertension (*iAUC*_10_ = 0.79), obstructive sleep apnea (*iAUC*_5_ = 0.86), myocardial infarction (*iAUC*_15_ = 0.80), and stroke (*iAUC*_15_ = 0.78). We also show that these outcomes can be accurately predicted in independent cohorts, including CV disease death (*iAUC*_10_ = 0.79), coronary heart disease death (*iAUC*_10_ = 0.74), obstructive sleep apnea (*iAUC*_5_ = 0.77), and stroke (*iAUC*_10_ = 0.60). We report increased performance across all outcomes compared to other sleep foundation models, such as SleepFM. Through correlational analyses and explainability approaches, we illustrate features most informative for risk at different horizons. We further demonstrate SleepJEPA can effectively estimate sleep stages with high accuracy (F1 = 0.77 [95% CI: 0.76 - 0.77]), objective daytime sleepiness with modest performance (AUC = 0.64 [0.57 - 0.71]), and type 1 narcolepsy (AUC = 0.88 [0.68 - 0.97]), allowing for comprehensive labeling and disease risk assessment from hPSG signals.

## 1 Introduction

Sleep disturbances are a global epidemic with deleterious effects (Canever et al., 2024; Chattu et al., 2018a,b; Lim et al., 2023; McWhorter et al., 2019; Melaku et al., 2019). During the COVID-19 pandemic, it was estimated that 40.49% of the global population have sleep disturbances (Jahrami et al., 2022), with an estimated 10% increase during the pandemic (Partinen et al., 2021). Sleep disorders constitute a substantial portion of these estimates, with global prevalence of insomnia, obstructive sleep apnea (OSA), and restless leg syndrome affecting 850 million, 1 billion, and 325 million, respectively (Benjafield et al., 2025, 2019; Song et al.). Further, there is evidence that sleep disturbances are increasing due to continued social and economic changes, an aging population, smartphone use, and climate change (Canever et al., 2024; Chattu et al., 2018a; Dissing et al., 2021; Lechat et al., 2025; Minor et al., 2022). Compounding this issue, sleep disturbances are well known to be linked to widespread conditions such as cardiovascular (CV), metabolic, and neurological disease (Chattu et al., 2018b; Lim et al., 2023). Thus, identifying the links between sleep and disease is essential.

While in-lab polysomnography (PSG) (Type I) is the clinical benchmark for identifying sleep problems, its reliance on manual scoring and resource-heavy infrastructure has limited its scalability (Chinoy et al., 2020; Goldstein et al., 2020; Jafari and Mohsenin, 2010; Malhotra et al.). Consequently, clinical practice has pivoted toward simplified home testing (Type III) (Braun et al., 2024a; Kundel and Shah, 2017), which, while convenient, sacrifices the rich physiological signals found in electroencephalograms (EEG), electrooculograms (EOG), and electromyograms (EMG). This creates a diagnostic “blind spot” where potentially predictive markers of cardiovascular, metabolic, and neurological risk — often embedded in sleep architecture — are lost. Historically, the acquisition of high-fidelity Type II hPSG has been largely confined to research due to the technical complexity of self-application and the prohibitive cost of both the device and manual scoring. However, rapid advancements in wearable sensor technology are now making EEG, EOG, and EMG data increasingly viable in the home environment (Vitazkova et al., 2025), reducing barriers to access while maintaining accuracy compared to in-lab studies (Braun et al., 2024b; Korkalainen et al., 2021). Despite these advances, the link between sleep problems measured using hPSG and risk for future disease is not well established due to the complexity and size of sleep study signals, reliance on standard or aggregated PSG metrics, and studies focusing on either in-lab PSG signals or an expanded set that is not generalizable to hPSG.

Artificial intelligence (AI) can bridge the gap between raw sleep study signals and disease risk. Traditionally, AI approaches in sleep have assisted in labeling tasks including sleep staging and respiratory events (Lee et al., 2025; Nasiri et al., 2025; Nassi et al., 2022; Olesen et al., 2021; Perslev et al., 2021; Phan et al., 2023, 2022a,b; Zan and Yildiz, 2023), with the goal of reducing variability in scoring and removing access bottlenecks to trained sleep professionals (Nasiri et al., 2025). To better understand the relationship between sleep and future risk for disease, research has also shown that signal features extracted from sleep studies with AI algorithms can estimate CV disease and mortality risk (Ambale-Venkatesh et al., 2017; Butler et al., 2019; Cavaillès et al., 2025; Koscova et al., 2024; Zhang et al., 2020), cognitive impairment (Cavaillès et al., 2025; Haghayegh et al., 2025; Sun et al., 2025), age (Brink-Kjaer et al., 2022), and narcolepsy (Stephansen et al., 2018). However, these works utilize single modality or aggregated sleep study signal features. This could limit the vast data streams of the polysomnogram, where potentially subtle features or dynamics across multiple channels could be predictive of long-term disease risk, such as in the EEG (Lechat et al., 2022a,b). Also, these studies primarily assess performance on single cohort datasets, which limits interpretations for out-of-distribution performance. Finally, models focus on single outcomes for prediction using supervised techniques, despite the multitude of risks associated with sleep disturbances.

Recent advances in AI, especially self-supervision, have shown promise in learning from multimodal data, including images and text for multi-outcome risk estimation with impressive performance on out-of-distribution data (Christensen et al., 2024; Huang et al., 2023; Ma et al., 2025; Xiang et al., 2025; Xu et al., 2024; Zhang et al., 2024; Zhou et al., 2023). In the sleep domain, self-supervised models on raw polysomnogram data have good performance for sleep staging prediction and disease risk estimation (Banville et al., 2021; Fox et al., 2025a; He et al., 2025; Ogg and Coon, 2024; Thapa et al., 2026; Weng et al., 2025). However, training techniques for sleep foundational models, such as SleepFM (Thapa et al., 2026), lag vision models and rely on masked autoencoding or contrastive learning. Newer methods, such as joint embedding predictive architectures (JEPA) that learn representations in the latent space via predicting masked inputs from context input, could reduce learning signal channel noise and increase learning of physiological states during pretraining, compared to masked autoencoding or contrastive learning (Assran et al., 2023; Kim, 2024; Weimann and Conrad, 2025).

Here we develop and validate SleepJEPA, the first JEPA sleep signal representation model, that learns features of multichannel, variable length (6-12 hours) hPSG signal channels, providing a scalable framework for identifying systemic disease risk. Our goals for this work were to (1) build a JEPA signal model with full night (6-12 hour) hPSG data, (2) show that full night representations contain meaningful features of sleep physiology for sleep staging, daytime sleepiness, and age prediction tasks, (3) examine representation performance on estimating long-term (up to 15 year) disease risk, including CV, metabolic, and neurological conditions on held-out and independent test sets, and (4) identify clinically meaningful features underlying risk estimation results.

## 2 Methods

### 2.1 SleepJEPA Architecture

JEPA is a self-supervised training framework that learns representations via three key networks – a context encoder, target encoder, and predictor. First, randomly selected tokens of input data (or patches of signal data) are selected as targets (e.g., what the model will “predict” during self-supervised training). The remaining tokens (or a subset of them) are used as context (e.g., the initial input to the self-supervised model). Second, context tokens are passed through the context encoder network to create context embeddings. Learnable mask tokens are appended to the context to specify which targets to predict. These embeddings are then processed through a smaller predictor network. Third, the target embeddings are generated by passing the full, non-masked sample through the target encoder, which is a copy of the context encoder with exponentially weighted moving average weight adjustments. The target embeddings are then subset to the originally selected targets and compared to the predictor’s output via a regression loss function. This embedding space prediction is hypothesized to learn more semantic representations (Assran et al., 2023), while also reducing noisy reconstructions and representational collapse (Jing et al., 2022).

SleepJEPA incorporates long signal data from sleep studies. It aims to reduce learning signal noise common with masked autoencoding self-supervised learning approaches and focus on learning representations of the physiological state of sleep in the latent space. Previous works demonstrated that the JEPA architecture with signal data improves performance over other self-supervised approaches in ECG classification (Kim, 2024; Weimann and Conrad, 2025), wearables (Xie et al., 2025), and ICU risk prediction (Fox et al., 2025b). SleepJEPA is based on the PhysioJEPA architecture published by our group (Fox et al., 2025b), which utilizes patch time series transformers (Nie et al., 2023) as the context, target, and predictor networks. An overview of the context encoder, target encoder, and predictor are detailed in Extended Data Figures 1, 2, and 3. SleepJEPA uses signals from seven signal channels, including a single EEG channel (C4-M1 or C3-M2), left EOG (E1-M2), chin EMG, augmented lead II ECG, SpO_2_, and respiratory inductance plethysmography (RIP) signals across the thorax and abdomen, of any length (6 to 12 hours) to accommodate varying length sleep studies. For datasets, preprocessing, and model architecture specifics, see the Supplementary Methods.

#### 2.1.1 Self-Supervised Training Monitoring

During self-supervised learning, it is difficult to assess when representations are robust and have avoided dimensional or complete collapse. This is a notable problem in contrastive learning approaches (Jing et al., 2022; Tian et al., 2021) and JEPA architectures (Assran et al., 2023; Balestriero and LeCun, 2025). Thus, our model’s linear probing capabilities were evaluated with a validation set from the pretraining data at the end of every pretraining epoch. A logistic regression with 5-fold cross validation was fit to estimate sleep stages, including wake, stage 1 (N1), stage 2 (N2), stage 3 (N3), and rapid eye movement (REM) sleep. The pretraining epoch with the best performance was selected for finetuning of all downstream tasks.

#### 2.1.2 Assessing Representations

After representation learning, finetuning heads were trained with frozen representations to estimate sleep stages, age, and daytime sleepiness to assess representation features and their associations with outcomes and labels. For sleep staging, a 2-layer bidirectional gated recurrent unit (GRU) was utilized as previously described (Fox et al., 2025a). See the Supplementary Methods for datasets, preprocessing, and model architecture specifics.

Age was estimated using an attentive regression model. See Extended Data Figure 4 for an illustration and the Supplementary Methods for training and dataset specifics.

Finally, to assess daytime sleepiness, the overnight PSG from the preceding night of a mean sleep latency test (MSLT) was utilized to estimate an MSLT score of less than or equal to 8 minutes, an indicator for objective daytime sleepiness (Goldbart et al., 2014). Additionally, narcolepsy outcomes were also used to train another model to estimate type 1 narcolepsy. Narcolepsy labels reflect mean sleep latency of less than or equal to 8 minutes and presence of at least 2 sleep onset REM periods (REM latency of less than or equal to 15 minutes in at least 2 naps) (Stephansen et al., 2018). For both daytime sleepiness outcomes, an attentive classifier head was utilized. Additional training and dataset details are provided in the Supplementary Methods.

### 2.2 Estimating Long-term Disease Risk

Ten outcomes spanning CV, metabolic, and neurological conditions were evaluated. Independent finetuning heads were trained per outcome. Details on the datasets and outcome processing are provided in the Supplementary Methods.

To estimate long-term disease risk, an attentive classifier was trained using a discrete hazard loss function. Discrete hazard loss functions estimate risk at multiple discrete times, handle non-proportional hazards, and optimize training in comparison to other hazard functions (Gensheimer and Narasimhan, 2019). Traditionally, Cox proportional hazard models, as used in SleepFM, are employed to estimate risk at a single time point in machine learning settings (Ching et al., 2018; Katzman et al., 2018; Thapa et al., 2026). Yet, in clinical scenarios, the proportional hazards assumption is more than likely violated as observation effects vary across time. Further, when training with Cox proportional hazard models, each sample’s estimated risk depends on all other samples’ risks that survived longer. Thus, when using gradient descent, the batch size influences the risk estimates, since individual batch-samples’ risks are only being compared to other batch samples and not all other samples in the training dataset. The discrete hazard loss function does not have this issue.

We explored three models per outcome: a model that utilizes only sleep representations (SleepJEPA), a model that includes age and sex hazards modeling with sleep representations (SleepJEPA+), and a baseline demographics (age and sex) only model. The third model is to elucidate the effects of demographic variables on predictions over time and better assess the performance of our sleep representations. See the Supplementary Methods for additional details on model architectures.

### 2.3 Model Evaluation

Models were evaluated with standard metrics depending on the task. Sleep stages were evaluated with area under the receiver operating characteristics curve (AUC), Cohen’s Kappa, F1, confusion matrices, and accuracy. Age estimation was assessed with mean absolute error and Pearson’s correlation coefficient. Daytime sleepiness was assessed with AUC, area under the precision recall curve (AUPRC), recall, and specificity. 95% bootstrapped confidence intervals (n = 999 resamples) were calculated to assess uncertainty.

For long-term disease outcomes, the cumulative/dynamic AUC (C/D AUC) score was used to assess performance at specific risk horizons (Lambert and Chevret, 2016). This score was also integrated (iAUC) to assess performance of previous risk horizons up to the current and serves as an aggregated metric of C/D AUC. Additionally, to assess calibration and discrimination, the integrated Brier score (IBS) was used to measure the model’s performance across all risk horizons. The IBS calculates the inverse probability of censoring weights (IPCW) to estimate the censoring distribution of the training dataset, providing a more reliable population estimate (Graf et al., 1999). Finally, the IPCW concordance index (C-Index) was used to evaluate performance at specific risk horizons (Uno et al., 2011). Again, the IPCW C-Index relies on the training dataset censoring distribution and provides a more reliable estimate, compared to using the testing dataset censoring distribution.

### 2.4 Clinical Interpretation

To better understand the predicted risk scores, the Pearson correlation was calculated between the predicted risk scores and baseline polysomnogram features, including age, ESS, BMI, stage 1, 2, 3, REM, and overall sleep time, sleep efficiency, average SpO_2_, minimum SpO_2_, obstructive apnea hypopnea index (AHI), and central AHI. Correlations are illustrated in clustered heatmaps to show features associated with risk predictions at different risk horizons and in box plots to show average correlation across all risk horizons. Correlation cutoffs were defined as weak (r = 0.1 to 0.3), moderate (r = 0.4 to 0.6), and strong (r = 0.7 to 1.0) (Akoglu, 2018). Additionally, survival curves were plotted to better understand the predicted risk quartiles for each outcome in both the held-out and independent test sets. Platt scaling was performed for calibration using the validation set for each model-outcome.

### 2.5 Model Explainability

The integrated gradients method (Sundararajan et al., 2017) was implemented to better understand representations in relation to estimating long-term disease risk. This method helps attribute an importance value to channel representations. Attributions for each 3-second patch were computed for each outcome to identify patches of signals and signal channels contributing to risk prediction results for outcomes from held-out test sets. Following, attributions were aggregated to sleep stage to identify sleep stages and signal channels contributing to predictions. Additional details regarding this method are described in the Supplementary Methods.

## 3 Results

SleepJEPA was pretrained with 34,291 sleep studies (n = 27,067 unique patients) and finetuned on 21,227 separate sleep studies (n = 18,839 unique patients). In total, our model was trained, validated, and tested on 422,035 hours of sleep study data with 7 signal channels. Demographics of the datasets used in pretraining and finetuning are reported in Table 1. The median age of our pretraining dataset was 55 (interquartile range [IQR]: 43 - 66), median BMI was 30.1 (26.0 - 35.5), median Epworth Sleepiness Score (ESS) was 8 (4 - 12), and 44.6% were female. The finetuning cohort had a median age of 68 (58 - 75), a median BMI of 27.7 (24.9 - 31.4), a median ESS of 7 (4 - 10), and 40.0% were female. An overview of the SleepJEPA design is in Figure 1.

**Figure 1:**
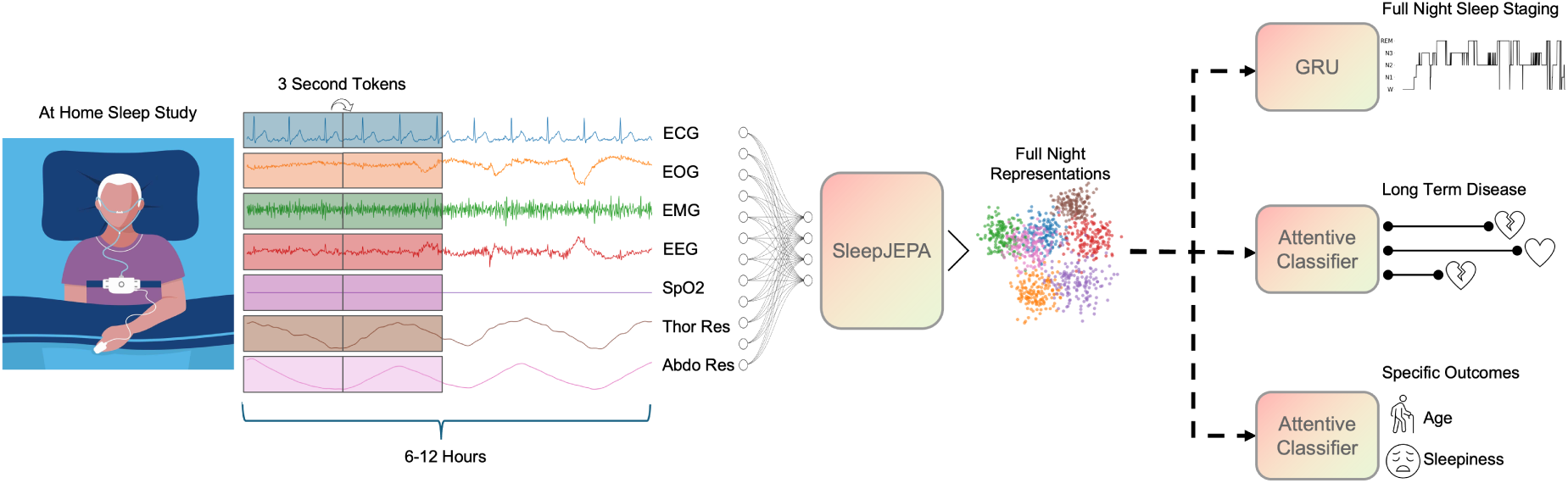
SleepJEPA Overview. Full night (6-12 hour) hPSG data from 7 channels are tokenized into 3 second patches and passed through SleepJEPA to generate representations. Representations are used as input to train additional models including sleep staging, long-term disease risk, and sleep study specific outcomes.

**Table 1:** Sleep Study Demographics, if available. MNC does not provide any demographic information. Apnea hypopnea index (AHI) represents all apneas independent of arousal and hypopneas with > 30% flow reduction and >= 3% oxygen desaturation or with arousal. ESS: Epworth Sleepiness Score.

|  | n Studies | n Patients | Median Age (IQR) | Sex, % Female | Race, % White | Median BMI (IQR) | Median ESS (IQR) | Median AHI (IQR) | Collection Period | Split |
| --- | --- | --- | --- | --- | --- | --- | --- | --- | --- | --- |
| Human Sleep Project | 22870 | 17697 | 55 (43-66) | 43.03% | - | 30.38 (26.23-35.77) | 8 (4-12) | - | 2007-2023 | Pretrain |
| Mount Sinai PSG | 6728 | 6123 | 54 (40-65) | 50.71% | 37.22% | 29.53 (25.34-35.26) | 6 (1-11) | - | 2017-2024 | Pretrain |
| SHHS Visit 1 | 5755 | 5755 | 63 (55-72) | 52.38% | 84.57% | 27.49 (24.67-30.76) | 7 (4-11) | 13.21 (6.69-23.78) | 1995-1998 | Finetune |
| MrOS Visit 1 | 2898 | 2898 | 76 (72-80) | 0.00% | 90.72% | 26.72 (24.56-29.33) | 6 (3-8) | 17.10 (9.09-29.49) | 2005 | Finetune |
| SHHS Visit 2 | 2608 | 2608 | 67 (60-76) | 53.64% | 87.23% | 27.71 (24.86-31.13) | 7 (4-10) | 13.53 (6.73-24.50) | 2001-2003 | Finetune |
| WSC | 2522 | 1116 | 60 (54-66) | 46.39% | 95.84% | 30.30 (26.52-35.30) | 8 (5-11) | 14.20 (6.20-27.48) | 2000-2015 | Pretrain/Finetune |
| MESA | 2054 | 2054 | 68 (62-76) | 53.65% | 36.17% | 27.88 (24.63-31.83) | 5 (3-8) | 18.36 (9.49-33.23) | 2010-2012 | Finetune |
| MNC | 498 | 498 | - | - | - | - | - | - | - | Pretrain/Finetune |
| APPLES | 1055 | 1055 | 52 (43-61) | 34.60% | 76.30% | 30.62 (27.09-36.11) | 10 (7-13) | 34.20 (19.80-53.80) | 2003-2008 | Pretrain/Finetune |
| MrOS Visit 2 | 996 | 996 | 80 (77-84) | 0.00% | 87.35% | 27.00 (24.00-29.00) | 6 (4-9) | 17.00 (9.00-31.00) | 2009-2012 | Finetune |

After 16 epochs, SleepJEPA reached the best performance, based on the AUC score on sleep stage linear probing. Training loss decreased and validation AUC increased up to this point, after which training loss dropped substantially while validation AUC began to plateau. While validation sleep stage AUC scores were optimal (> 0.70) at lower training losses, epoch 16 was chosen as it provides the best linearly separable representations and could help avoid representational collapse in later epochs. See Supplementary Figure 1 for training loss and validation AUC curves.

### 3.1 SleepJEPA Performance in Sleep Staging

Sleep staging distributions are available in Supplementary Table 1. On a held-out test set of 2,531 sleep studies, SleepJEPA representations achieved a macro-AUC of 0.97 (95% CI: 0.97 - 0.97), with a Cohen’s Kappa of 0.80 (0.80 - 0.81), a macro F1 score of 0.77 (0.76 - 0.77), and a micro accuracy of 0.86 (0.86 - 0.86). The model was independently tested on 4,050 sleep studies, reaching a macro-AUC score of 0.92 (95% CI: 0.91 - 0.92), Cohen’s Kappa was 0.66 (0.65 - 0.66), macro F1 was 0.61 (0.61 - 0.62), and micro accuracy was 0.77 (0.77 - 0.77). Confusion matrices are reported in Figure 2.

**Figure 2:**
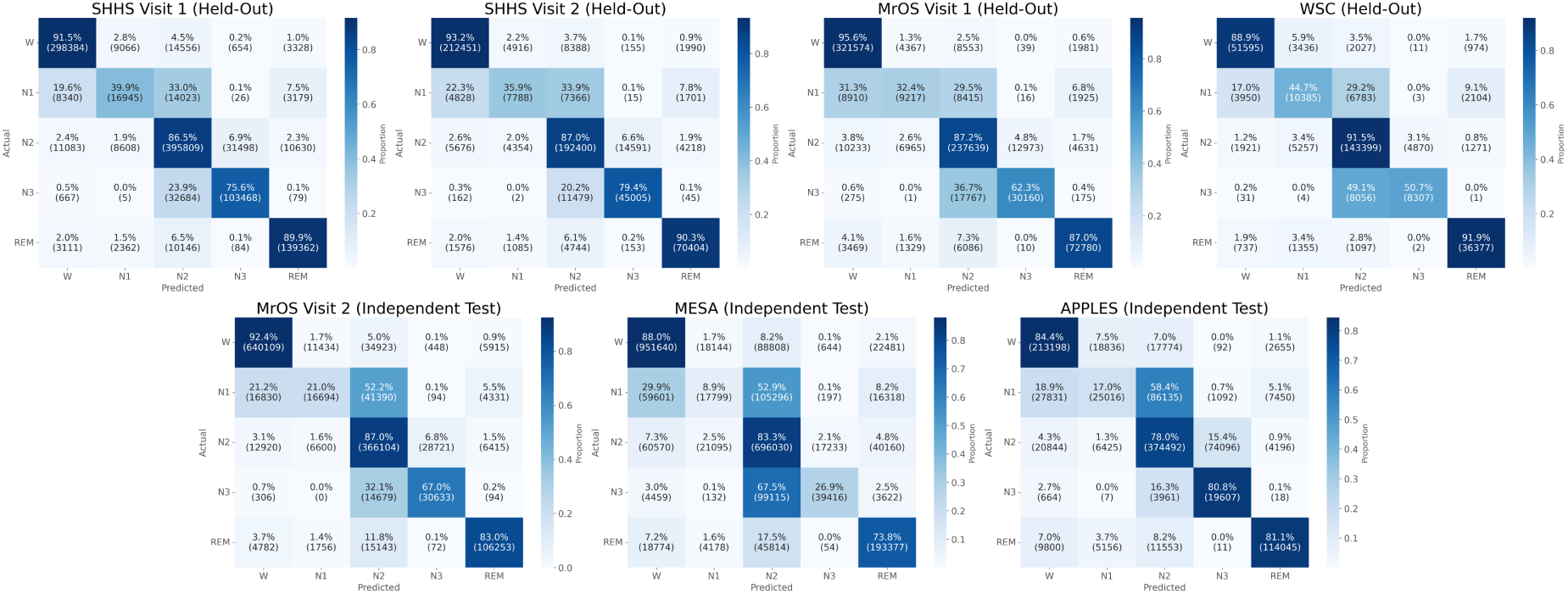
SleepJEPA Sleep Stage Confusion Matrices. Row normalized confusion matrices (with counts) for sleep stage classification on held-out test sets SHHS Visit 1, SHHS Visit 2, MrOS Visit 1, and WSC (top row) and independent test sets MrOS Visit 2, MESA, and APPLES (bottom row).

Compared to other state-of-the-art sleep staging models, SleepJEPA performs comparably. L-SeqSleepNet (Phan et al., 2023), SleepTransformer (Phan et al., 2022a), XSleepNet (Phan et al., 2022b), USleep (Perslev et al., 2021), FullSleepNet (Zan and Yildiz, 2023), and SleepXViT (Lee et al., 2025) had the highest performance based on macro F1 scores on the Sleep Heart Health Study (SHHS) (macro F1 = 80.0-81.0), compared to SleepJEPA (77.2). However, these models were all trained end-to-end for the sleep staging task. On the Osteoporotic Fractures in Men Study (MrOS), USleep and SleepFM (Thapa et al., 2026) achieved the highest macro F1 scores on a held-out test set, reaching 77.0 for USleep and 75.0 for SleepFM. SleepJEPA achieved a macro F1 score of 74.3 on only MrOS Visit 1, with fewer signal channels. On the Wisconsin Sleep Cohort (WSC), SleepJEPA and PFTSleep (Fox et al., 2025a) achieve the highest performance with 74.7 and 73.8 macro F1 scores, respectively.

On independent testing sets, SleepJEPA was compared to PFTSleep, a patch foundational transformer model trained via a masked autoencoder framework that similarly learn representations to finetune for sleep staging. SleepJEPA modestly outperforms PFTSleep across the MrOS Visit 2, the Multi-Ethnic Study of Atherosclerosis (MESA), and the Apnea Positive Pressure Long-term Efficacy Study (APPLES) test sets, reaching macro F1 scores of 69.4, 57.2, and 60.10, compared to PFTSleep’s scores of 66.7, 56.7, and 58.5, respectively. SleepFM evaluated independent sleep staging performance on small cohorts of 255 patients from the Danish Center for Sleep Medicine (DCSM) sleep staging dataset and 151 patients from the Haaglanden Medisch Centrum (HMC) sleep staging database, achieving macro F1 scores of 0.68 (for DCSM) and 0.55 (for HMC) (Thapa et al., 2026). SleepJEPA independently evaluated 4,050 sleep studies across MrOS visit 2, MESA, and APPLES. Additional results comparing models by dataset are reported in Table 2.

**Table 2:** Sleep Stage Predictive Performance Comparison. Table adapted from Phan et al. (Phan et al., 2023) and Fox et al. (Fox et al., 2025a). Bold, underlined indicates best performance and bold only indicates second best performance. Held-out test sets were not identical. * Indicates the datasets are independent test sets.

| Dataset | Model | Micro Accuracy | Cohen's Kappa | Macro F1 | F1 (W) | F1 (N1) | F1 (N2) | F1 (N3) | F1 (REM) |
| --- | --- | --- | --- | --- | --- | --- | --- | --- | --- |
| SHHS | SleepJEPa | 85.98 | 0.8 | 77.21 | 92.87 | 41.45 | 85.84 | 76.29 | 89.65 |
|  | SleepFM (Thapa et al., 2026) | - | - | 78 | 92 | 49 | 84 | 72 | **91** |
|  | PFTSleep (Fox et al., 2025a) | 86.07 | 0.8 | 76.53 | 92.17 | 35.92 | 86.47 | 78.27 | 89.82 |
|  | SleepXViT (Visit 1) (Lee et al., 2025) | **88.07** | **0.83** | 80.18 | **93.29** | 45.34 | **88.57** | 83.5 | 90.18 |
|  | L-SeqSleepNet (Phan et al., 2023) | 87.6 | **0.83** | 80.3 | 92.4 | 48.6 | 88.2 | 83.9 | 88.5 |
|  | SeqSleepNet (Phan et al., 2019) | 87.2 | **0.82** | 80.2 | 91.8 | 49.1 | 88.2 | 83.5 | 88.2 |
|  | SleepTransformer (Phan et al., 2022a) | **87.7** | **0.83** | 80.1 | 92.2 | 46.1 | 88.3 | **85.2** | 88.6 |
|  | XSleepNet1 (Phan et al., 2022b) | 87.6 | **0.83** | **80.7** | 91.6 | **51.4** | **88.5** | 85 | 88.4 |
|  | XSleepNet2 (Phan et al., 2022b) | 87.5 | **0.83** | **81** | 92 | 49.9 | 88.3 | 85 | 88.2 |
|  | U-Sleep (Perslev et al., 2021) | - | - | 80 | 93 | **51** | 87 | 76 | **92** |
|  | FullSleepNet (Zan and Yildiz, 2023) | 87.55 | **0.83** | 80.3 | 92.8 | 48.5 | 88 | 82.4 | 89.8 |
|  | Olesen et al. (Olesen et al., 2021) | 87.1 | **0.82** | 78.8 | **94.1** | 47.8 | 87.9 | 74.3 | 89.9 |
|  | AttnSleep (Eldele et al., 2021) | 84.2 | 0.78 | 75.3 | 86.7 | 33.2 | 87.1 | **87.1** | 82.1 |
| MrOS | SleepJEPa (Visit 1) | **87.25** | **0.81** | 74.26 | **94.45** | 36.6 | 86.27 | 65.87 | 88.13 |
|  | SleepFM (Thapa et al., 2026) | - | - | 75 | 94 | **41** | 86 | 65 | **90** |
|  | PFTSleep (Visit 1) (Fox et al., 2025a) | **87.96** | **0.82** | 72.57 | **95.03** | 23.1 | **86.76** | **69.65** | **88.31** |
|  | Olesen et al. (Olesen et al., 2021) | 84.3 | 0.76 | - | - | - | - | - | - |
|  | USleep (Perslev et al., 2021) | - | - | 77 | 93 | **46** | **87** | **68** | 88 |
| WSC | SleepJEPa | **85.07** | **0.77** | **74.65** | 88.74 | **47.57** | **90.17** | **56.14** | **90.61** |
|  | PFTSleep (Fox et al., 2025a) | **85.47** | **0.78** | **73.77** | **90.03** | 33.83 | 90.05 | **65.24** | 89.69 |
|  | Olesen et al. (Olesen et al., 2021) | 82.6 | 0.72 | - | - | - | - | - | - |
|  | Olesen et al. 2018 (Olesen et al., 2018) | 85 | 0.75 | 72.6 | **88.8** | **49.7** | **90.2** | 47.3 | 87.2 |
| MrOS Visit 2* | SleepJEPa | **84.86** | **0.76** | **69.41** | **93.6** | **28.83** | **81.99** | 57.97 | **84.66** |
|  | PFTSleep (Fox et al., 2025a) | 84.36 | 0.75 | 66.72 | 92.86 | 13 | 81.93 | **61.56** | 84.22 |
| MESA* | SleepJEPa | **75.18** | **0.63** | **57.2** | **87.44** | **13.66** | 74.44 | 38.59 | 71.87 |
|  | PFTSleep (Fox et al., 2025a) | 72.72 | 0.6 | 56.73 | 79.79 | 3.11 | **76.03** | **47.34** | **77.39** |
| APPLES* | SleepJEPa | 71.42 | 0.59 | **60.1** | 81.23 | **24.65** | **76.9** | **32.91** | 84.81 |
|  | PFTSleep (Fox et al., 2025a) | **71.79** | 0.59 | 58.47 | **84.59** | 11.83 | 76.12 | 32.42 | **87.38** |

### 3.2 Association of SleepJEPA with Chronological Age

To understand the relationship between SleepJEPA’s representations and age, an attentive regression model was trained to predict chronological age. On the held-out test set of 1,728 studies, comprising MrOS Visit 1 and SHHS Visit 1 cohorts, mean absolute error (MAE) was 5.51 and the Pearson correlation was 0.80. For our independent test set on 4,576 studies, comprising MESA and WSC cohorts, the MAE was 6.47, and Pearson correlation was 0.61. MAE by age is shown in Figure 3 and depicts performance across ages, related to the distribution of the test sets. Generally, SleepJEPA performs the best at predicting age for those in the 55-70 range, while younger and older predictions become more variable. Predicted vs actual age plots are shown in Supplementary Figure 2 for both the held-out and independent test sets.

**Figure 3:**
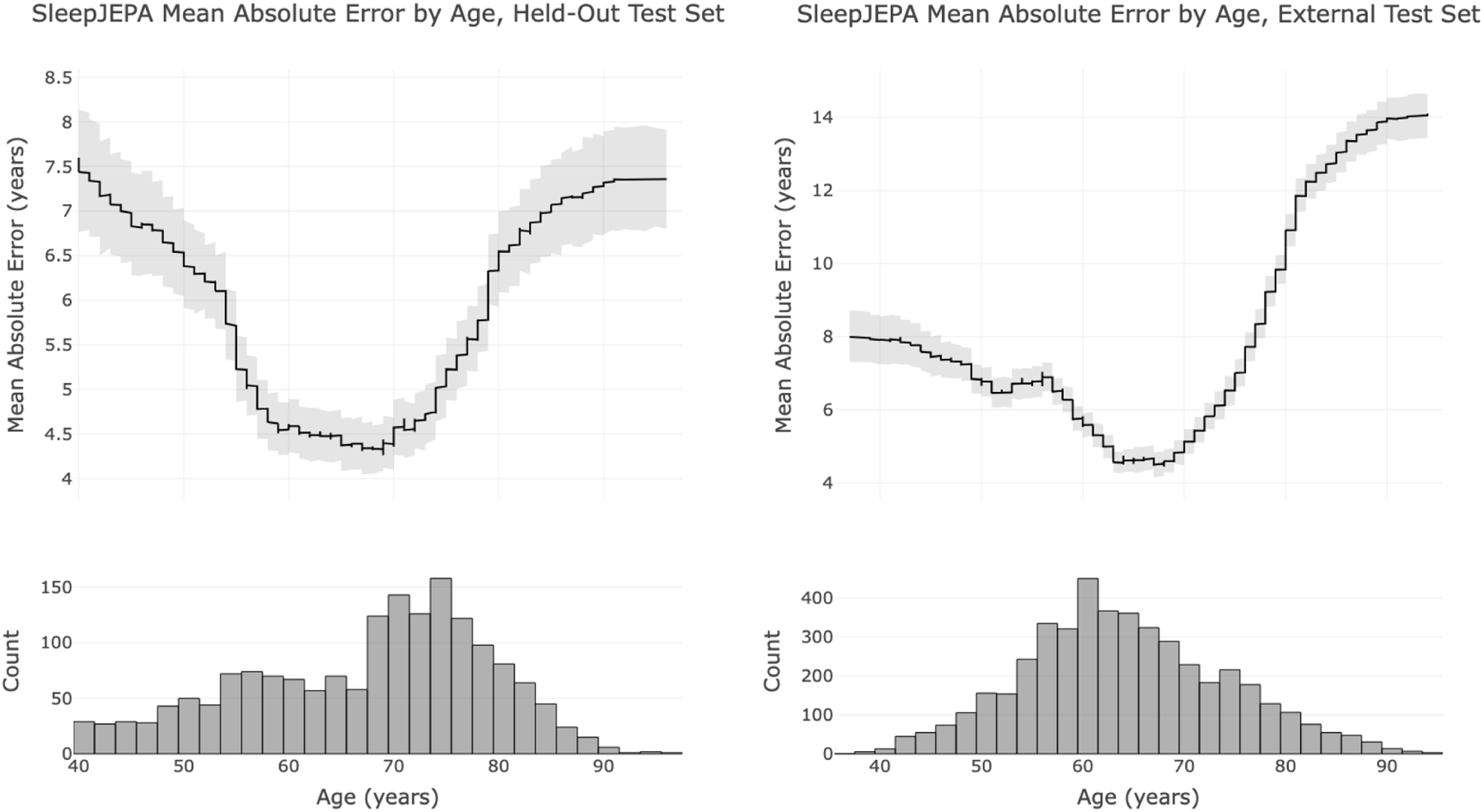
SleepJEPA Age Predictive Performance. Smoothed mean absolute error by age with 95% bootstrapped confidence intervals (n = 999 resamples) for the held-out test (left) and independent test set (right).

### 3.3 Association of SleepJEPA with Daytime Sleepiness

To assess objective daytime sleepiness and type 1 narcolepsy status, two additional attentive classifiers were trained for each outcome. Outcome distributions are reported in Supplementary Table 2. For type 1 narcolepsy prediction, SleepJEPA representations reached an AUC score of 0.88 (95% CI: 0.68 - 0.97) and AUPRC of 0.53 (0.15 - 0.79) compared to the positive prevalence of 8.8%. When independently testing the MSLT outcomes from WSC, the model had reduced performance (AUC = 0.56 [0.52 - 0.59] and AUPRC = 0.30 [0.26 - 0.34]). When predicting objective daytime sleepiness based on MSLT scores, SleepJEPA AUC scores were 0.64 (0.57 - 0.71), with an AUPRC of 0.36 (0.26 - 0.44) compared to the positive prevalence of 25.4%. When independently testing the Mignot Nature Communications (MNC) narcolepsy outcomes, AUC was 0.65 (0.58 - 0.73), and AUPRC was 0.14 (0.09 - 0.19). Additional metrics are reported in Table 3.

**Table 3:** SleepJEPA Daytime Sleepiness Outcomes Predictive Performance. SleepJEPA was trained to estimate type 1 narcolepsy with the MNC cohort and objective daytime sleepiness (MSLT <= 8) on the WSC cohort. Each model was then used to estimate the other’s outcome as an independent test. Performance metrics are shown above for the held-out test sets (same dataset as training) and independent test set for both models. 95% bootstrapped confidence intervals (n = 999 resamples) are reported.

| Outcomes | Test Set | AUC | Average Precision | Recall | Specificity |
| --- | --- | --- | --- | --- | --- |
| Objective Daytime Sleepiness | Held-Out | 0.64 (0.57 - 0.71) | 0.36 (0.26 - 0.44) | 0.70 (0.60 - 0.80) | 0.50 (0.43 - 0.56) |
|  | Independent (MNC, Type 1 Narcolepsy) | 0.65 (0.58 - 0.73) | 0.14 (0.09 - 0.19) | 0.36 (0.23 - 0.51) | 0.78 (0.75 - 0.82) |
| Type 1 Narcolepsy | Held-Out | 0.88 (0.68 - 0.97) | 0.53 (0.15 - 0.79) | 0.30 (0.08 - 0.75) | 0.98 (0.92 - 1.00) |
| | Independent (WSC, MSLT ≤ 8) | 0.56 (0.52 - 0.59) | 0.30 (0.26 - 0.34) | 0.00 (0.00 - 0.01) | 0.99 (0.99 - 1.00) |

### 3.4 SleepJEPA Risk Estimation for Long-term Disease

SleepJEPA representations were frozen and used as input into 10 separate attentive classifiers to estimate long-term disease risk (SleepJEPA). Dataset event distributions are in Extended Data Table 1, and training, validation, and testing splits are in Supplementary Table 3. Each model was trained for 10 epochs for lightweight finetuning to disease outcomes. Additionally, a baseline demographics model (age and sex) and a SleepJEPA representations model with age and sex hazards modeling (SleepJEPA+) were trained for each outcome. Training details are provided in the Supplementary Methods. The iAUC, C/D AUC, and IPCW C-Index were calculated at 1, 2, 3, 5, 10, and 15 years and reported in Table 4. Additionally, C/D AUC curves over 15 years for all outcomes are illustrated in Figure 4.

**Figure 4:**
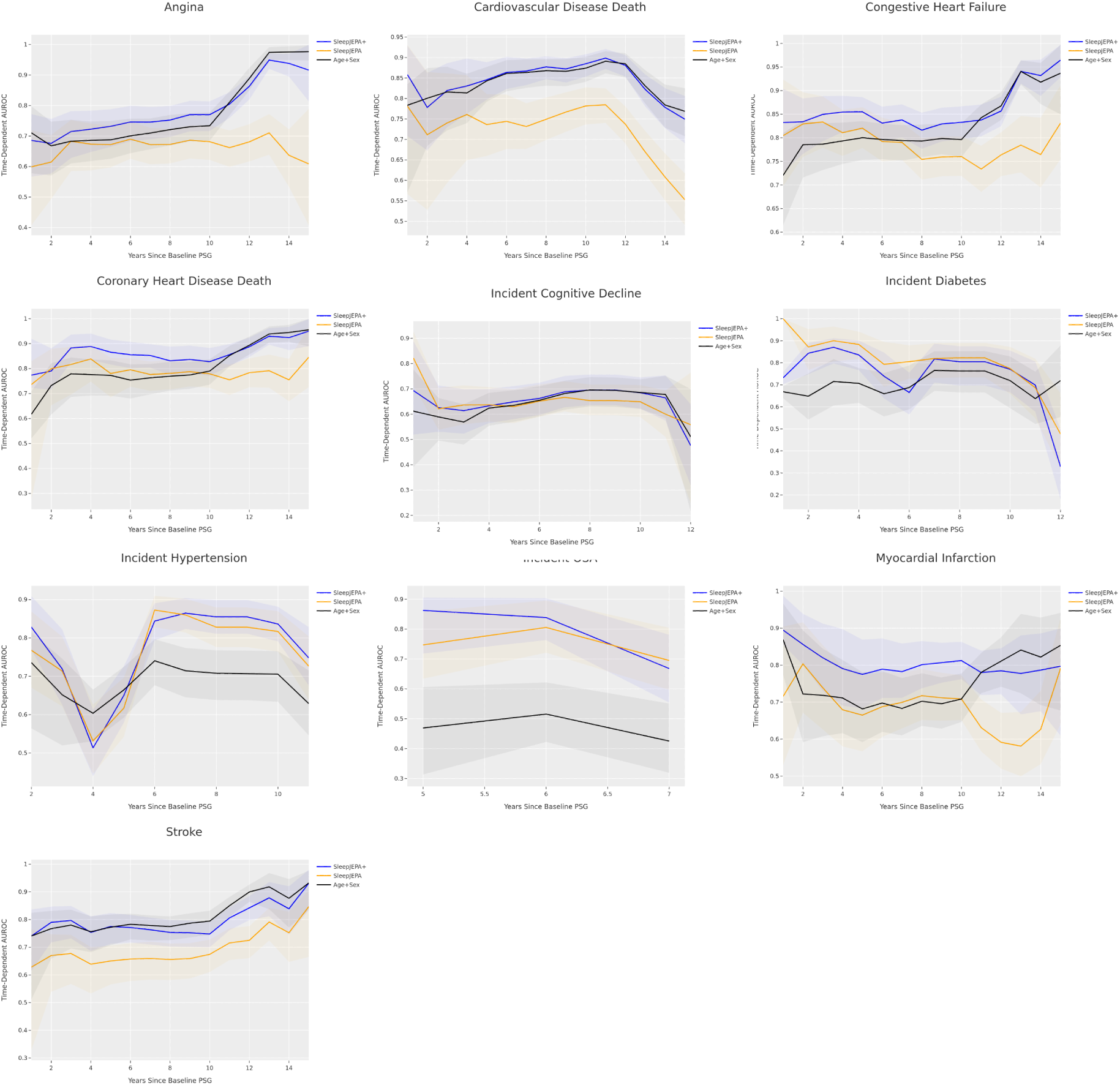
C/D AUC plots for each outcome on the 20% held-out test sets, up to 15 years. SleepJEPA+, SleepJEPA, and the baseline demographics model are reported. 95% bootstrapped confidence intervals (n = 999 resamples) are shaded for each model.

**Table 4:** SleepJEPA long-term outcomes predictive performance at different forecast lengths. SleepJEPA+, SleepJEPA, and the baseline demographics model are reported. iAUC is the integral of the C/D AUC, up to that year. C-Index is the IPCW C-Index for the reported year. Bold, underlined indicates best performance and bold indicates second best performance. Results are for the 20% held-out test set for each outcome. 95% bootstrapped confidence intervals (n = 999 resamples) are reported for C/D AUC and C-Index.

| Outcome | Year | iAUC<br>SleepJEPa+ | SleepJEPa | Age+Sex | C/D AUROC<br>SleepJEPa+ | SleepJEPa | Age+Sex | C-Index<br>SleepJEPa+ | SleepJEPa | Age+Sex |
| --- | --- | --- | --- | --- | --- | --- | --- | --- | --- | --- |
| Angina | 1 | **0.69** | 0.60 | **0.71** | **0.69 (0.57-0.77)** | 0.60 (0.41-0.72) | **0.71 (0.58-0.80)** | **0.68 (0.62-0.73)** | 0.60 (0.53-0.65) | **0.71 (0.65-0.76)** |
|  | 2 | **0.68** | 0.61 | **0.69** | **0.68 (0.58-0.76)** | 0.61 (0.49-0.70) | **0.67 (0.57-0.75)** | **0.67 (0.61-0.72)** | 0.61 (0.55-0.66) | **0.66 (0.61-0.72)** |
|  | 3 | **0.69** | 0.63 | **0.69** | **0.71 (0.63-0.78)** | **0.68 (0.58-0.75)** | **0.68 (0.61-0.75)** | **0.71 (0.65-0.76)** | **0.68 (0.62-0.73)** | **0.68 (0.61-0.73)** |
|  | 5 | **0.70** | 0.64 | **0.69** | **0.73 (0.66-0.79)** | 0.67 (0.60-0.73) | **0.69 (0.63-0.75)** | **0.72 (0.65-0.78)** | **0.67 (0.59-0.73)** | **0.67 (0.60-0.74)** |
|  | 10 | **0.72** | 0.66 | **0.70** | **0.77 (0.72-0.81)** | 0.68 (0.62-0.74) | **0.73 (0.68-0.78)** | **0.73 (0.64-0.81)** | 0.66 (0.57-0.74) | **0.69 (0.59-0.77)** |
|  | 15 | **0.73** | 0.66 | **0.71** | **0.92 (0.81-1.00)** | 0.61 (0.41-0.79) | **0.98 (0.95-1.00)** | **0.74 (0.45-0.91)** | 0.65 (0.37-0.85) | **0.71 (0.42-0.89)** |
| Cardiovascular Disease Death | 1 | **0.86** | 0.78 | **0.78** | **0.86 (0.71-0.93)** | **0.78 (0.57-0.93)** | **0.78 (0.57-0.84)** | **0.86 (0.80-0.90)** | **0.78 (0.71-0.84)** | **0.78 (0.71-0.84)** |
|  | 2 | **0.80** | 0.73 | **0.80** | **0.78 (0.67-0.86)** | 0.71 (0.53-0.86) | **0.80 (0.68-0.87)** | **0.78 (0.72-0.83)** | 0.71 (0.65-0.77) | **0.80 (0.74-0.85)** |
|  | 3 | **0.81** | 0.73 | **0.80** | **0.82 (0.73-0.89)** | 0.74 (0.59-0.86) | **0.82 (0.72-0.88)** | **0.81 (0.75-0.86)** | 0.74 (0.67-0.80) | **0.81 (0.75-0.86)** |
|  | 5 | **0.83** | 0.74 | **0.82** | **0.85 (0.80-0.89)** | 0.74 (0.66-0.81) | **0.84 (0.79-0.88)** | **0.83 (0.78-0.88)** | 0.73 (0.67-0.79) | **0.83 (0.77-0.88)** |
|  | 10 | **0.86** | 0.75 | **0.85** | **0.88 (0.85-0.91)** | 0.78 (0.74-0.83) | **0.87 (0.84-0.90)** | **0.85 (0.79-0.90)** | 0.76 (0.69-0.81) | **0.84 (0.78-0.89)** |
|  | 15 | **0.83** | 0.68 | **0.83** | **0.75 (0.69-0.81)** | 0.55 (0.49-0.62) | **0.77 (0.71-0.83)** | **0.80 (0.70-0.87)** | 0.66 (0.55-0.76) | **0.80 (0.70-0.88)** |
| Congestive Heart Failure | 1 | **0.83** | **0.80** | 0.72 | **0.83 (0.73-0.90)** | **0.80 (0.70-0.92)** | 0.72 (0.61-0.81) | **0.83 (0.79-0.86)** | **0.80 (0.76-0.84)** | 0.72 (0.67-0.76) |
|  | 2 | **0.83** | **0.82** | 0.75 | **0.83 (0.77-0.88)** | **0.83 (0.76-0.90)** | 0.79 (0.72-0.84) | **0.83 (0.78-0.86)** | **0.82 (0.78-0.86)** | 0.78 (0.73-0.82) |
|  | 3 | **0.84** | **0.82** | 0.76 | **0.85 (0.79-0.89)** | **0.83 (0.78-0.89)** | 0.79 (0.73-0.83) | **0.84 (0.79-0.88)** | **0.83 (0.78-0.86)** | 0.78 (0.73-0.82) |
|  | 5 | **0.84** | **0.82** | 0.78 | **0.86 (0.82-0.89)** | **0.82 (0.78-0.86)** | 0.80 (0.75-0.84) | **0.84 (0.79-0.88)** | **0.81 (0.76-0.85)** | 0.78 (0.73-0.83) |
|  | 10 | **0.84** | **0.80** | 0.79 | **0.83 (0.80-0.87)** | 0.76 (0.72-0.80) | **0.80 (0.76-0.83)** | **0.80 (0.73-0.85)** | 0.73 (0.67-0.79) | **0.75 (0.69-0.81)** |
|  | 15 | **0.85** | 0.79 | **0.81** | **0.96 (0.93-1.00)** | 0.83 (0.75-0.91) | **0.94 (0.85-1.00)** | **0.80 (0.59-0.92)** | 0.71 (0.49-0.86) | **0.75 (0.53-0.89)** |
| Coronary Heart Disease Death | 1 | **0.77** | **0.74** | 0.62 | **0.77 (0.72-0.92)** | **0.74 (0.28-0.83)** | 0.62 (0.52-0.71) | **0.77 (0.70-0.84)** | **0.74 (0.66-0.80)** | 0.62 (0.53-0.70) |
|  | 2 | **0.79** | **0.78** | 0.70 | **0.79 (0.72-0.88)** | **0.80 (0.65-0.88)** | 0.73 (0.62-0.82) | **0.79 (0.72-0.84)** | **0.80 (0.73-0.85)** | 0.73 (0.66-0.79) |
|  | 3 | **0.82** | **0.79** | 0.73 | **0.88 (0.83-0.94)** | **0.82 (0.70-0.89)** | 0.78 (0.69-0.85) | **0.88 (0.82-0.92)** | **0.81 (0.74-0.87)** | 0.77 (0.70-0.83) |
|  | 5 | **0.84** | **0.79** | 0.74 | **0.87 (0.82-0.92)** | **0.78 (0.69-0.84)** | 0.77 (0.69-0.83) | **0.86 (0.79-0.91)** | **0.77 (0.70-0.84)** | 0.76 (0.68-0.83) |
|  | 10 | **0.84** | **0.79** | 0.76 | **0.83 (0.77-0.88)** | 0.78 (0.72-0.83) | **0.79 (0.74-0.84)** | **0.80 (0.71-0.87)** | **0.76 (0.67-0.84)** | 0.75 (0.65-0.82) |
|  | 15 | **0.85** | **0.78** | 0.78 | **0.95 (0.89-1.00)** | 0.85 (0.73-0.97) | **0.96 (0.89-1.00)** | **0.82 (0.47-0.96)** | 0.74 (0.40-0.92) | **0.75 (0.41-0.93)** |
| Incident Cognitive Decline | 1 | **0.69** | **0.82** | 0.61 | **0.69 (0.52-0.89)** | **0.82 (0.71-0.93)** | 0.61 (0.39-0.78) | **0.69 (0.59-0.77)** | **0.82 (0.73-0.88)** | 0.61 (0.51-0.70) |
|  | 2 | **0.64** | **0.66** | 0.59 | **0.63 (0.53-0.71)** | **0.62 (0.54-0.70)** | 0.59 (0.50-0.66) | **0.62 (0.53-0.69)** | **0.62 (0.53-0.69)** | 0.58 (0.50-0.66) |
|  | 3 | **0.64** | **0.65** | 0.59 | **0.61 (0.52-0.70)** | **0.64 (0.56-0.71)** | 0.57 (0.48-0.64) | **0.60 (0.51-0.69)** | **0.63 (0.54-0.72)** | 0.56 (0.47-0.65) |
|  | 5 | **0.64** | **0.65** | 0.61 | **0.65 (0.58-0.71)** | 0.63 (0.56-0.70) | **0.64 (0.57-0.69)** | **0.61 (0.50-0.71)** | **0.61 (0.49-0.71)** | 0.59 (0.48-0.70) |
|  | 10 | **0.65** | **0.65** | 0.63 | **0.68 (0.62-0.75)** | 0.65 (0.59-0.71) | **0.69 (0.62-0.74)** | **0.63 (0.51-0.74)** | **0.62 (0.49-0.72)** | **0.62 (0.49-0.73)** |
| Incident Diabetes | 1 | **0.73** | **1.00** | 0.67 | **0.73 (0.70-0.76)** | **1.00 (1.00-1.00)** | 0.67 (0.64-0.69) | **0.73 (0.62-0.81)** | **1.00 (0.95-1.00)** | 0.67 (0.56-0.76) |
|  | 2 | **0.83** | **0.89** | 0.65 | **0.84 (0.75-0.89)** | **0.87 (0.77-0.95)** | 0.65 (0.54-0.75) | **0.84 (0.76-0.90)** | **0.87 (0.79-0.92)** | 0.65 (0.55-0.73) |
|  | 3 | **0.84** | **0.89** | 0.66 | **0.87 (0.80-0.91)** | **0.90 (0.81-0.96)** | 0.72 (0.61-0.82) | **0.86 (0.78-0.92)** | **0.89 (0.81-0.94)** | 0.71 (0.61-0.80) |
|  | 5 | **0.81** | **0.87** | 0.68 | **0.74 (0.64-0.85)** | **0.79 (0.69-0.90)** | 0.66 (0.56-0.75) | **0.72 (0.55-0.84)** | **0.78 (0.62-0.89)** | 0.65 (0.48-0.79) |
|  | 10 | **0.72** | **0.82** | 0.69 | **0.77 (0.67-0.86)** | **0.77 (0.66-0.87)** | 0.72 (0.59-0.83) | **0.63 (0.44-0.79)** | **0.64 (0.45-0.80)** | 0.58 (0.39-0.75) |
| Incident Hypertension | 2 | **0.83** | **0.77** | 0.74 | **0.83 (0.77-0.91)** | **0.77 (0.67-0.87)** | 0.74 (0.56-0.81) | **0.82 (0.72-0.89)** | **0.76 (0.66-0.84)** | 0.73 (0.62-0.82) |
|  | 3 | **0.77** | **0.74** | 0.69 | **0.72 (0.63-0.82)** | **0.71 (0.62-0.80)** | 0.65 (0.52-0.75) | **0.71 (0.60-0.80)** | **0.71 (0.60-0.80)** | 0.65 (0.53-0.75) |
|  | 5 | **0.65** | 0.63 | **0.65** | **0.65 (0.58-0.73)** | 0.62 (0.54-0.70) | **0.66 (0.59-0.72)** | **0.64 (0.53-0.73)** | 0.62 (0.51-0.72) | **0.65 (0.54-0.74)** |
|  | 10 | **0.78** | **0.79** | 0.71 | **0.84 (0.79-0.88)** | **0.82 (0.76-0.87)** | 0.71 (0.63-0.77) | **0.70 (0.59-0.79)** | **0.68 (0.57-0.77)** | 0.60 (0.49-0.70) |
| Incident OSA | 5 | **0.86** | **0.75** | 0.47 | **0.86 (0.72-0.91)** | **0.75 (0.63-0.86)** | 0.47 (0.31-0.61) | **0.86 (0.72-0.93)** | **0.75 (0.60-0.85)** | 0.47 (0.33-0.62) |
| Myocardial Infarction | 1 | **0.89** | 0.72 | **0.87** | **0.89 (0.86-0.99)** | 0.72 (0.53-0.90) | **0.87 (0.77-0.97)** | **0.89 (0.83-0.94)** | 0.71 (0.63-0.79) | **0.87 (0.79-0.92)** |
|  | 2 | **0.87** | **0.77** | 0.77 | **0.86 (0.80-0.94)** | **0.80 (0.67-0.92)** | 0.72 (0.59-0.89) | **0.85 (0.78-0.90)** | **0.80 (0.73-0.86)** | 0.72 (0.64-0.79) |
|  | 3 | **0.85** | **0.76** | 0.75 | **0.82 (0.73-0.91)** | **0.74 (0.63-0.85)** | 0.72 (0.61-0.84) | **0.81 (0.74-0.87)** | **0.74 (0.66-0.81)** | 0.71 (0.63-0.78) |
|  | 5 | **0.82** | 0.72 | **0.73** | **0.77 (0.68-0.87)** | 0.66 (0.57-0.77) | **0.68 (0.59-0.78)** | **0.77 (0.68-0.83)** | 0.66 (0.58-0.74) | **0.67 (0.58-0.75)** |
|  | 10 | **0.81** | **0.71** | 0.71 | **0.81 (0.76-0.86)** | **0.71 (0.65-0.77)** | **0.71 (0.65-0.78)** | **0.80 (0.70-0.87)** | **0.71 (0.61-0.79)** | 0.69 (0.58-0.77) |
|  | 15 | **0.80** | 0.68 | **0.74** | **0.80 (0.61-0.90)** | 0.79 (0.64-0.93) | **0.85 (0.68-0.94)** | **0.74 (0.42-0.92)** | **0.65 (0.34-0.87)** | **0.65 (0.34-0.87)** |
| Stroke | 1 | **0.74** | 0.63 | **0.74** | **0.74 (0.61-0.84)** | 0.63 (0.33-0.76) | **0.74 (0.51-0.82)** | **0.74 (0.68-0.79)** | 0.63 (0.56-0.69) | **0.74 (0.68-0.79)** |
|  | 2 | **0.77** | 0.66 | **0.76** | **0.79 (0.72-0.85)** | 0.67 (0.54-0.75) | **0.77 (0.66-0.83)** | **0.79 (0.74-0.83)** | 0.67 (0.61-0.72) | **0.76 (0.71-0.81)** |
|  | 3 | **0.78** | 0.66 | **0.76** | **0.80 (0.73-0.85)** | 0.68 (0.57-0.74) | **0.78 (0.70-0.84)** | **0.79 (0.74-0.84)** | 0.67 (0.61-0.73) | **0.77 (0.72-0.82)** |
|  | 5 | **0.77** | 0.65 | **0.76** | **0.78 (0.71-0.82)** | 0.65 (0.57-0.71) | **0.77 (0.71-0.82)** | **0.76 (0.70-0.81)** | 0.64 (0.57-0.70) | **0.76 (0.69-0.81)** |
|  | 10 | **0.77** | 0.66 | **0.77** | **0.75 (0.70-0.79)** | 0.67 (0.61-0.73) | **0.79 (0.75-0.83)** | **0.71 (0.63-0.78)** | 0.65 (0.56-0.73) | **0.75 (0.67-0.82)** |
|  | 15 | **0.78** | 0.67 | **0.79** | **0.93 (0.83-0.98)** | 0.85 (0.67-0.91) | **0.93 (0.84-0.98)** | **0.69 (0.42-0.88)** | 0.65 (0.38-0.85) | **0.73 (0.45-0.90)** |

Across nearly all time horizons and outcomes, SleepJEPA+ improved predictive performance compared to the demographics baseline. As forecasting horizons extend further from the baseline sleep study, participants are naturally older, making age an increasingly strong predictor for nearly all outcomes. Consequently, SleepJEPA+ or SleepJEPA performed better at closer horizons compared to the demographics only baseline, with top iAUC scores at 1-, 2-, 3-, and 5-year forecasts, except for angina at 1-and 2- year risk. At 10 and 15-year forecasts, SleepJEPA+ outperforms the demographics baseline for angina, congestive heart failure, coronary heart disease death, incident cognitive decline, diabetes, hypertension, and myocardial infarction. The demographics baseline performed comparably for estimating CV disease death and stroke at 10- and 15-year forecasts. IBS is reported in Supplementary Table 4, and indicated all models were calibrated well, with similar scores.

Independent test sets were evaluated for outcomes with additional data. On the MESA study for CV disease death, SleepJEPA+ outperforms the baseline demographics at all risk horizons, with a 10-year iAUC score of 0.79 compared to 0.64 for SleepJEPA and 0.73 for the baseline demographics model. Coronary heart disease death was also evaluated in the MESA study. SleepJEPA+ had a 10-year iAUC score of 0.74, compared to SleepJEPA (0.53) and the demographics baseline (0.65). OSA was assessed using the WSC dataset. SleepJEPA+ and SleepJEPA outperformed the demographics baseline at all years evaluated. For myocardial infarction and stroke on the MESA dataset, SleepJEPA+ and SleepJEPA appear to lose predictive power and perform worse than the demographics baseline, except for stroke at 10 years. While the demographics baseline performs better, especially at 1-year for myocardial infarction (iAUC = 0.69), all other horizons are below 0.60. C/D AUC curves for independent testing are illustrated in Extended Data Figure 5, and iAUC, C/D AUC, and IPCW C-Index are reported in Table 5. IBS scores are reported in Supplementary Table 5.

**Table 5:**
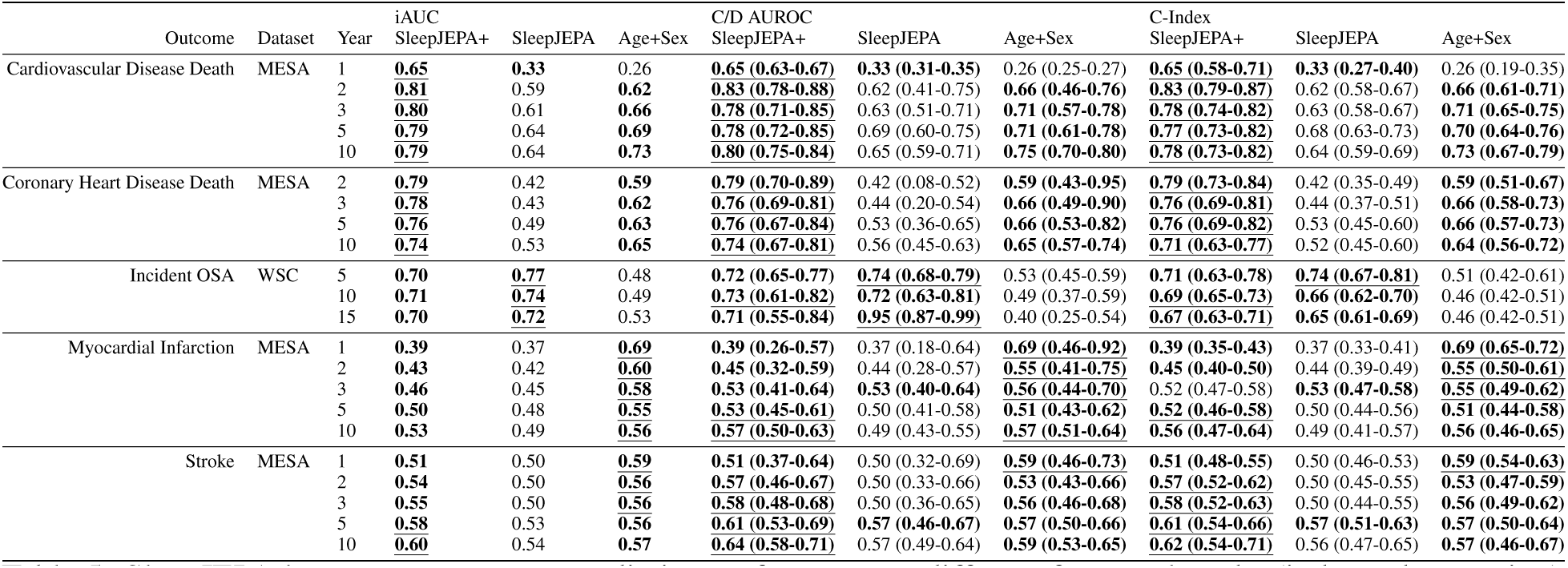
SleepJEPA long-term outcomes predictive performance at different forecast lengths (independent testing). SleepJEPA+, SleepJEPA, and the baseline demographics model are reported. iAUC is the integral of the C/D AUC, up to that year. C-Index is the IPCW C-Index for the reported year. Bold, underlined indicates best performance and bold indicates second best performance. Results are for the independent test sets for available outcomes. 95% bootstrapped confidence intervals (n = 999 resamples) are reported for C/D AUC and C-Index.

Compared to SleepFM (Thapa et al., 2026) risk predictions at 6-years and He et al. (He et al., 2025) risk predictions at 5-years, SleepJEPA+ improves predictive performance for all outcomes, including angina, CV disease death, congestive heart failure, coronary heart disease death, incident hypertension, myocardial infarction, and stroke in a held-out SHHS dataset. Results are reported in Table 6.

**Table 6:** SleepJEPA long-term outcomes predictive performance compared to other models on held-out SHHS. SleepJEPA+ is compared to SleepFM (Thapa et al., 2026) at 6 years with AUC and C-Index metrics. SleepJEPA+ is also compared to He et al. at 5 years with AUC (He et al., 2025). Models reported all included demographics. Bold indicates the best performance. 95% bootstrapped confidence intervals (n = 999 resamples) are reported for AUC and C-Index (if available).

| Outcome | 6-Year AUROC<br>SleepJEPa+ | SleepFM | 6-Year C-Index<br>SleepJEPa+ | SleepFM | 5-Year AUROC<br>SleepJEPa+ | CV Outcome (HE) |
| --- | --- | --- | --- | --- | --- | --- |
| Angina | **0.83 (0.77-0.88)** | 0.76 (0.72-0.81) | **0.80 (0.72-0.87)** | 0.73 (0.71-0.75) | 0.82 (0.76-0.88) | - |
| Cardiovascular Disease Death | **0.90 (0.85-0.95)** | 0.88 (0.83-0.91) | **0.89 (0.81-0.94)** | 0.86 (0.83-0.89) | **0.89 (0.84-0.94)** | 0.783 |
| Congestive Heart Failure | **0.88 (0.84-0.92)** | 0.85 (0.82-0.88) | **0.86 (0.80-0.90)** | 0.83 (0.81-0.86) | **0.89 (0.86-0.93)** | 0.848 |
| Coronary Heart Disease Death | **0.88 (0.83-0.93)** | 0.87 (0.82-0.92) | **0.87 (0.80-0.92)** | 0.86 (0.82-0.89) | 0.89 (0.85-0.94) | - |
| Incident Hypertension | 0.73 (0.63-0.90) | - | 0.59 (0.43-0.74) | - | **0.76 (0.69-0.84)** | 0.597 |
| Myocardial Infarction | **0.79 (0.72-0.86)** | 0.78 (0.71-0.84) | **0.77 (0.69-0.84)** | 0.75 (0.71-0.79) | **0.79 (0.71-0.87)** | 0.711 |
| Stroke | **0.83 (0.75-0.87)** | 0.82 (0.76-0.87) | **0.81 (0.74-0.87)** | **0.81 (0.76-0.85)** | 0.83 (0.75-0.88) | - |

### 3.5 Correlations of Sleep Summary Measures with SleepJEPA Risk Estimates

To further understand outcome predictions, cumulative risk scores from SleepJEPA across 15 years were extracted for the SHHS held-out dataset or MrOS held-out test set (for cognitive decline). Cumulative risk scores at 1, 2, 3, 5, 10, and 15 were correlated with common polysomnogram features for angina, CV disease death, congestive heart failure, coronary heart disease death, cognitive decline, diabetes, hypertension, OSA, myocardial infarction, and stroke. Mean correlation results are reported in Figure 5, and per year heatmaps are shown in Supplementary Figure 3.

Across all outcomes, age had a weak positive correlation (r = 0.10 to 0.39) with risk predictions and was most correlated with congestive heart failure risk predictions (r = 0.39 [95% CI: 0.37 - 0.40]). Sleep efficiency had weak negative correlations with all outcomes, with a maximum for CV disease death. Average or minimum SpO_2_ was also weakly negatively correlated for all outcomes except for cognitive decline, with a maximum average SpO_2_ for incident and maximum minimum SpO_2_ for CV disease death. Consequently, obstructive and central AHI were weakly positively correlated with risk predictions for all outcomes except cognitive decline, with a maximum for obstructive and central AHI for OSA. Arousal index followed a similar trend as obstructive and central AHI; however, it was not correlated with incident hypertension. REM sleep time was weakly negatively correlated for all outcomes, except incident diabetes, hypertension, and myocardial infarction. N1 sleep time was weakly positively correlated with CV disease death, congestive heart failure, coronary heart disease death, and diabetes with a maximum for CV disease death. N2 sleep time was weakly positively correlated with congestive heart failure, coronary heart disease death, and myocardial infarction. N3 sleep time was weakly negatively correlated with incident diabetes and myocardial infarction. Total sleep time was weakly negatively correlated with all outcomes, except for myocardial infarction. BMI was weakly positively correlated with incident diabetes, hypertension, and OSA, but not with other outcomes. ESS was not correlated with any outcome. See Supplementary Table 6 for all mean cumulative risk prediction correlations.

### 3.6 Survival modeling with SleepJEPA risk estimates

Survival curves for SleepJEPA+ for each outcome were plotted using the SHHS and MrOS held-out test sets. Results are shown in Figure 6. Independent test set results are shown in Extended Data Figure 6. SleepJEPA+ identifies quartiles of high, medium-high, medium-low, and low risk individuals at a 10-year horizon with modest separation for angina, CV disease death, congestive heart failure, coronary heart disease death, incident cognitive decline, myocardial infarction, and stroke. Other incident outcomes had more overlapping quartile confidence intervals. On the independent test set, risk quartiles were maintained for CV disease death and coronary heart disease death. Additionally, risk quartile separation was notably better for OSA compared to the held-out test set, likely due to the wider distribution of time to events in the WSC dataset. For myocardial infarction and stroke, the low- and high-risk quartiles were more clearly separated for stroke, with more overlap for the middle quartiles. For myocardial infarction, there was more overlap at shorter horizons across all quartiles, with more separation at later horizons.

### 3.7 Model Explainability

The integrated gradients method on SleepJEPA representations traced which representations and their corresponding sleep stage contribute to predictions for each outcome. Using samples from the SHHS held-out test set and MrOS (for incident cognitive decline), attribution scores were calculated and visualized in Supplementary Figures 4-13. For angina, per channel aggregated sleep stage attributions showed that ECG and EEG channels had the largest channel attributions, especially during REM and N3 sleep for ECG and N1, N2, and N3 for EEG. For CV disease death, ECG, EOG, respiratory, and SpO_2_ channels were the largest contributors especially while awake. Other sleep stages had similar attribution proportions. For congestive heart failure, the channel attributions from EEG’s wake sleep stage, SpO_2_ REM, and chin EMG N3 were highest. Coronary heart disease death had the largest attributions from thoracic RIP, ECG, and SpO_2_ with proportional contributions from each sleep stage. Incident cognitive decline had high attribution scores for respiratory channels, ECG, and EOG, with N2 and N3 sleep being the primary contributors. Notably, EOG N3 sleep appears to be a large contributor, compared to other sleep stages, for incident cognitive decline risk prediction. For incident diabetes, chin EMG wake, N1, and N2 stages and EOG wake, N2, and N3 had the largest attributions scores. Incident hypertension had large attributions from abdominal RIP, chin EMG, and SpO_2_ channels, with higher contributions from N3 sleep. For OSA, chin EMG and respiratory channels had the largest attribution scores, with larger proportions while awake and in REM sleep. Myocardial infarction had large attributions from ECG, EEG, and thoracic RIP, especially during REM and N3 sleep. Finally, for stroke, all channels had higher attribution spread proportionately across all stages.

**Figure 5:**
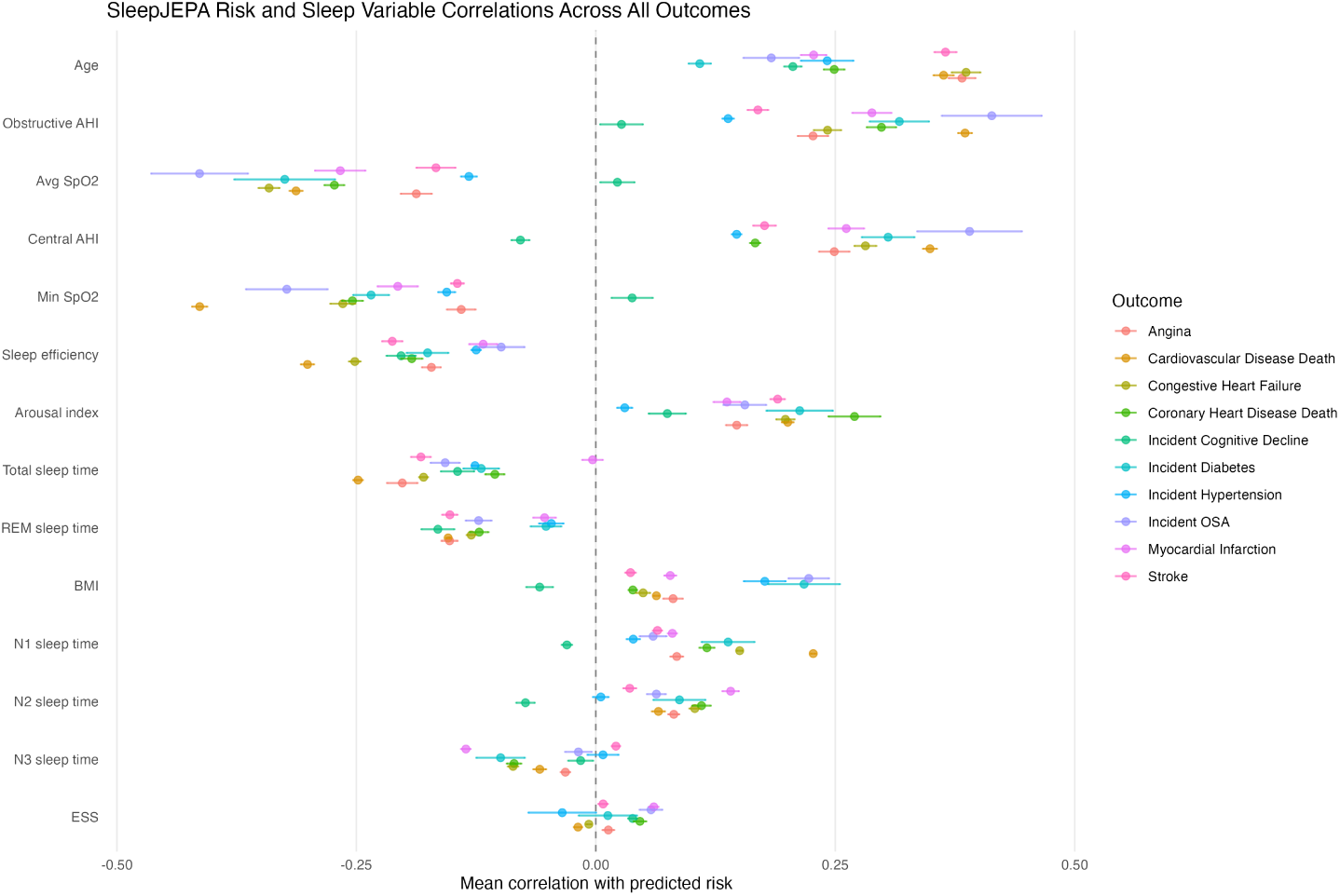
SleepJEPA representations predicted mean cumulative risk correlations across risk horizons (up to 15 years). The average Pearson correlation was calculated across available time horizons with normalized demographic and baseline PSG derived features. Correlations for each outcome are shown with 95% confidence intervals for the SHHS held-out test set and MrOS held-out test set (for cognitive decline).

While attributions may appear focused on specific channels, all outcomes have modest attributions across all signal channels and sleep stages. This may reflect SleepJEPA’s encoder attention mechanism, which processes channels independently using learned channel-agnostic attention weights, potentially diluting channel-specific information in the representation space and distributing attribution across channels and time.

## 4 Discussion

We present SleepJEPA, the first joint embedding predictive architecture that learns foundational representations of at-home PSG signal data in the latent space. SleepJEPA was trained, validated, and tested on 422,035 hours of sleep signals from 55,518 studies. Notably, SleepJEPA can encode hPSG recordings of variable length (between 6 and 12 hours), enabling scalability and versatility for use in research, clinical, and home settings.

SleepJEPA’s largest contribution is examining the relationship between sleep measured with at-home signals and long-term disease risk. While two prior works, including SleepFM, have applied self-supervised learning of PSG signals for related tasks (He et al., 2025; Thapa et al., 2026), SleepJEPA outperforms both models in all disease prediction tasks and further advances the field by building the first JEPA sleep signal model with only at-home sleep signals, independently training disease outcomes with a discrete hazard risk model for up to 15 years using the latent representations from JEPA training, testing disease outcomes on independent datasets, and examining performance in relation to clinical features, survival, and model explainability.

He et al. built a multichannel self-supervised model with at-home signal channels (ECG, EEG, and respiratory signals) to contrast CV disease outcome embeddings for risk estimation at 5 years using the SHHS dataset for training and WSC for independent validation (He et al., 2025). While He et al. independently builds logistic regression models for specific diseases, they fit a binary classification model per outcome and do not estimate risk with Cox proportional hazards or another survival method, potentially limiting performance, especially with censored data. Further, they limited their modeling to a 5-year risk horizon, which is insufficient for capturing the full spectrum of disease risk. Longer horizons are valuable for younger individuals with low short-term risk who may develop disease over time, while shorter horizons allow clinicians to identify older individuals at imminent risk.

**Figure 6:**
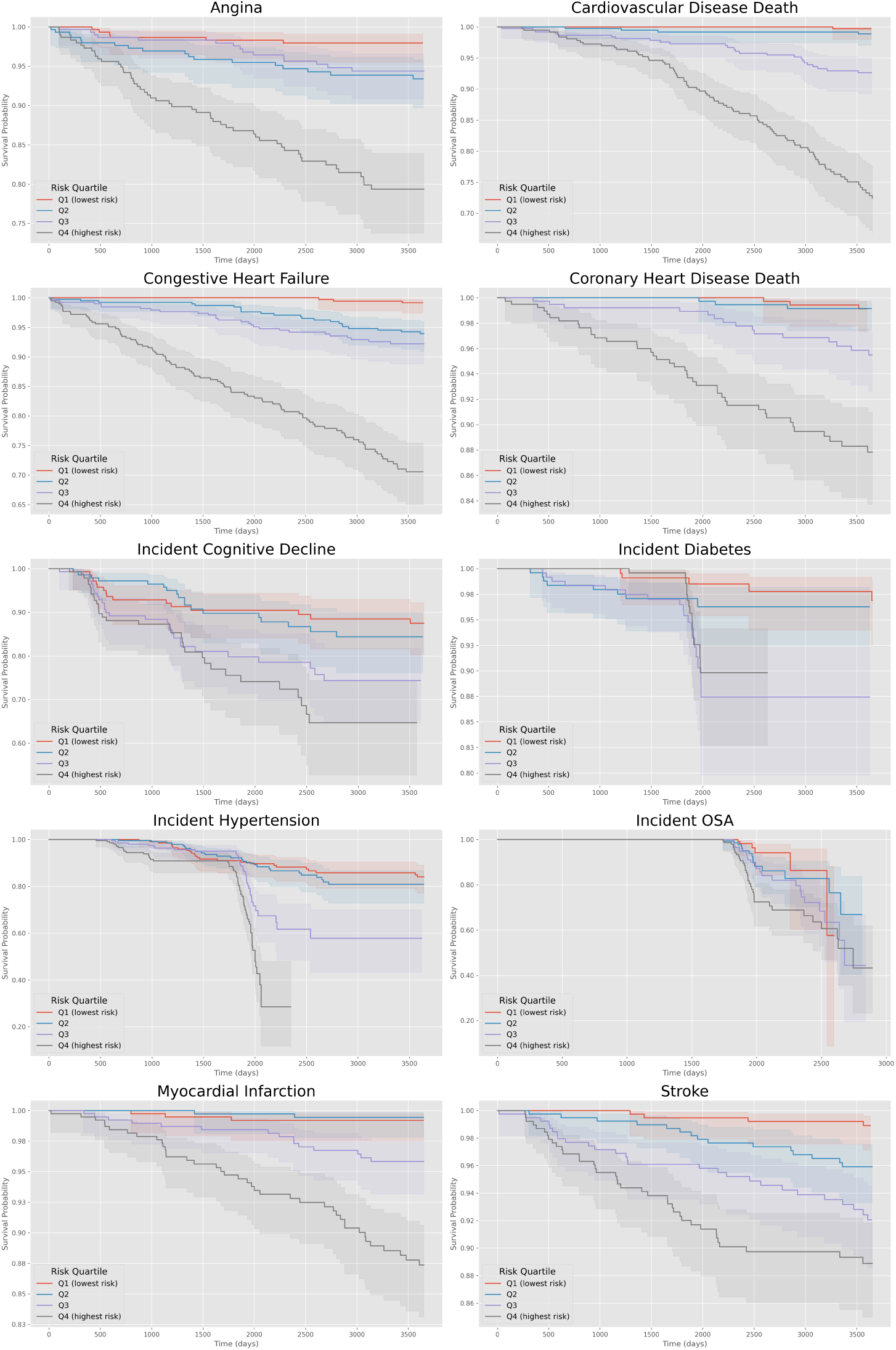
Survival curves at 10 years for all independent testing outcomes examining SleepJEPA+ risk quartiles on the MESA or WSC (for incident OSA) independent test sets. Probabilities were calibrated using Platt scaling per model-outcome using the validation set prior to survival monitoring. 95% confidence intervals are displayed in bands.

Thapa et al. built SleepFM, a multichannel self-supervised model with in-lab PSG brain, cardiac, and respiratory channels and showed that signal representations were predictive of age, sex, sleep staging, and a multilabel condition risk estimation task at 6 years with 1,041 disease outcomes (Thapa et al., 2026). They showed that these representations could be fine-tuned to the SHHS disease outcome dataset with impressive performance, despite only pretraining for one epoch. Notably, SleepFM was not evaluated on independent test sets, limiting assessment of generalizability. As with He et al., risk estimation was confined to a 6-year horizon, which may be insufficient for capturing long-term disease trajectories across diverse age groups. SleepFM predicts risk with a multilabel Cox proportional hazards model trained to estimate 1,041 disease at once; however, this multilabel task makes it difficult to determine which observations are predictive of labels given that patients present with multiple comorbidities and could confound results (Park et al., 2020). Additionally, as mentioned in the Methods, Cox proportional hazards modeling requires large batch sizes and proportional hazards, both of which were not discussed in SleepFM, especially when age effects vary over time.

SleepJEPA overcomes these limitations and illustrates the relationship between at-home sleep study signals and disease risk over time. SleepJEPA representations are more predictive at shorter time horizons, and age and sex hazard modeling enhances predictions for longer risk horizons. Additionally, SleepJEPA was independently tested on out of distribution datasets and maintained impressive performance in CV disease death, coronary heart disease death, and incident OSA risk predictions at various horizons. For myocardial infarction and stroke, performance decreased, potentially due to differences in labeling definitions compared to the training set, prevalence, or time to event distributions (see Supplementary Table 3).

Across all outcomes, sleep disruptions and length are related to a multitude of conditions, including the ones examined in this study (Yang et al., 2024). Sleep feature correlations and sleep stage attributions likely reflect these disruptions. However, identifying sleep features and attributions related to future disease via risk estimation helps detect potentially relevant clinical factors.

For CV outcomes, ECG channel attributions were higher for angina (ECG channel highest sleep stage attributions: REM and N3 sleep), CV disease death (awake and N1), coronary heart disease death (N1 and N2), congestive heart failure (awake and N1), myocardial infarction (REM and N2), and stroke (even across all stages). This expands upon previous work that showed ECG signals combined with sleep stages can predict risk for myocardial infarction, stroke, and chronic heart failure (Manimaran et al., 2025) by highlighting specific sleep stages contributing to predictions from the ECG signal channel.

SleepJEPA estimates for CV and coronary heart disease death and incident diabetes additionally revealed positive correlations between N1 sleep time, AHI, and arousal index, with high attributions when awake, potentially indicating disrupted sleep, a known risk factor for CV disease, coronary heart disease, and mortality (Ai et al., 2024; Huang et al., 2025; Nagai et al., 2010). For congestive heart failure, estimates revealed correlations with higher AHI and lower SpO_2_ and attributions in EEG while awake and SpO_2_ in REM, supporting previous research that has found heart failure as the top comorbidity with sleep disordered breathing (Khayat et al., 2013). Angina estimates had correlations with higher AHI, lower SpO_2_, and decreases in REM and total sleep time with the highest attributions in EEG signals across N1, N2, and N3 sleep stages. Notably, both shorter sleep (< 7 hours) and longer sleep (> 8 hours) individuals have been found to have a higher risk for grade 2 angina, with atypical (right-sided neck, or epigastric pain) angina manifesting in those with shorter sleep at higher odds (Alhaque Roomi et al., 2025). Thus, our results suggest that angina was more likely related to shorter duration of sleep and atypical type.

SleepJEPA’s estimates for incident hypertension showed attributions primarily in awake and N3 sleep stages, with negative correlations with total sleep time, likely indicating shorter sleep duration. Attributions in N3 sleep were interesting and could be related to a decreased stage 3 sleep proportion, which has been shown to increase risk for incident hypertension using SHHS (Javaheri et al., 2017). For incident cognitive decline, risk estimates highlighted respiratory channels in attributions, with smaller attributions in the SpO_2_ signal. Hypoxemia has been studied extensively in relation to cognitive decline (Blackwell et al., 2015; Yaffe et al., 2011). A recent work examined specifically that during REM sleep, hypoxemia is a strong link for cerebral small vessel disease and dementia (Berisha et al., 2025). However, attributions for SpO_2_ during REM sleep were low and correlations with oxygen saturation features were low. REM sleep time was negatively correlated with the predictions, possibly suggesting that our model was relying on more available features rather than REM SpO_2_, potentially in stage N2 and N3 where attribution proportions were highest.

For those with incident diabetes, SleepJEPA appears to be identifying disturbances in oxygen saturation given the negative correlation with SpO_2_ features; though, attribution scores for the SpO_2_ channel were lower compared to other channels. This aligns with a recent work that has shown hypoxemia during sleep increases blood glucose levels (SHINODA et al., 2018). Interestingly, EOG attributions were the 2nd highest contributing channel for diabetes risk predictions, especially in N3 sleep. While not traditionally thought of as distinguishing of sleep disorders, recent works have shown EOG’s ability to distinguish sleep stages (van Gorp et al., 2024) and identify sleep disorders (Jain and Ganesan, 2024). Further, a recent work has derived an EEG index of hyperglycemia (a coupling of NREM sleep spindles and slow oscillations) (R et al., 2023). Possibly, SleepJEPA is identifying related features in the EEG and EOG during N3 sleep when estimating diabetes risk.

Finally, SleepJEPA’s OSA risk estimations had expected attributions with EMG, respiratory, and SpO_2_ channels and correlations with decreased SpO_2_ and increased AHI. While awake attributions contained the highest proportions for ECG, EOG, EMG, and EEG, SpO_2_ had the highest proportion of attributions during REM sleep, potentially identifying clinically meaningful phenotypes for REM-related OSA risk (Verdi and Acıcan, 2025).

Unsurprisingly, the demographics baseline, which includes age and sex, performs strongly. Age is well known to be the main risk factor for common diseases (Niccoli and Partridge, 2012), and older people are more likely to have sleep problems and disorders (Madan Jha, 2023; Tatineny et al., 2020). Despite the high performance of the demographics baseline, it appears that modeling both sleep representations and age and sex hazards has an additive effect, perhaps to help extract more specific features from representations related to age and sex during finetuning. Interestingly, it also seems that the age only model overfits the training cohort data, given the discernible improvement of SleepJEPA in the independent testing evaluations; though, not necessarily for myocardial infarction or stroke risk estimations.

SleepJEPA also exhibits reliable performance in sleep staging, with modest improvement over a masked autoregression approach in N1 sleep and independent evaluation (PFTSleep (Fox et al., 2025a)). This highlights the advantage of learning representations in the latent space via JEPA. Further, SleepJEPA’s representations can estimate sleep stages with similar performance to other state-of-the-art models, while using frozen 3-second representations. While we aggregate to the traditional 30-second “sleep epoch” for comparison to other models, SleepJEPA’s 3-second representations — to our knowledge the smallest input size used for sleep staging — could capture more nuanced staging information and warrant further exploration. Finally, we extensively evaluate SleepJEPA representations for sleep stage classification on three large independent datasets. Performance remains high for wake, N2, and REM sleep and is especially compelling for MrOS visit 2, which had the highest macro F1 score (69.4) among the independent test sets. Given that SleepJEPA was finetuned with MrOS visit 1 for sleep staging, high performance on MrOS visit 2 suggests that SleepJEPA could be quickly finetuned to specific clinic data to support longitudinal patient monitoring and standardization of sleep technician labeling. Compared to SleepFM, both models have similar performance on held-out test sets; however, our evaluation on independent testing better illustrates performance variations by sleep stage and over time on 4,050 independent sleep studies. Further, SleepJEPA maintains performance across these independent sleep studies, while SleepFM has more varied performance across 406 sleep studies from two cohorts, a limitation mentioned in their manuscript (Thapa et al., 2026).

In addition to sleep staging, SleepJEPA’s representations appear to contain information related to daytime sleepiness and narcolepsy providing a relevant clinical metric without a high cost MSLT. SleepJEPA can estimate an MSLT score of 8 minutes or less (AUC = 0.64), an indicator for daytime sleepiness. When the daytime sleepiness model is independently tested to estimate type 1 narcolepsy, performance is similar (0.65), potentially indicating daytime sleepiness features are present in people with type 1 narcolepsy. When SleepJEPA’s representations are finetuned to estimate narcolepsy, performance suggests that there are even stronger features indicative of narcolepsy (0.88), but those features are not all present in those with daytime sleepiness (0.56). Recent work has shown that objective daytime sleepiness is associated with a higher risk of all-cause mortality (Dai et al., 2025). For patients with OSA, this could be related to the pulse oximetry signal (included in SleepJEPA), which has been shown to help distinguish those who have objective daytime sleepiness (Kainulainen et al., 2020).

Our study has limitations. While we do show that SleepJEPA excels across a multitude of horizons, conditions, and independent testing sets, our sleep study cohorts are primarily older individuals who presented with a sleep problem and were evaluated via polysomnography. The need for additional data across diverse cohorts is key to further sleep prognostic models. Second, incident outcome data labels were acquired from survey responses and other data collected at follow up visits potentially leading to less accurate labels or over-estimations of the time to event, which could imply performance is higher at longer risk horizons and lower at shorter risk horizons. Third, model explainability is inherently difficult with self-supervised deep learning approaches. While we kept signal channels separate during training and examined model attribution scores per-channel aggregated to sleep stage, identifying clinically interpretable features on signal data was not performed and should be a point of future work. Finally, differences in outcome labeling, prevalence, and time to event distributions for each dataset could lead to varied performance. This may be exemplified in our independent testing, where performance decreased, especially in myocardial infarction and stroke estimations.

Overall, SleepJEPA demonstrates that at-home sleep signals can serve as a scalable, non-invasive window into long-term disease risk. Integrated alongside other clinical risk assessment tools, SleepJEPA has the potential to provide meaningful insight into patients’ day to day sleep and long-term health. Future work should explore combining SleepJEPA with other clinical features to further improve predictive performance, and investigate whether reduced signal sets, such as Type III sleep signals and wearable devices, can maintain performance, lowering barriers to widespread clinical deployment.

## Data Availability

The Human Sleep Project is available at https://bdsp.io/content/hsp/2.0/. The Sleep Heart Health Study, the Wisconsin Sleep Cohort, the Osteoporotic Fractures in Men Study (MrOS), the Multi-Ethnic Study of Atherosclerosis (MESA), the Mignot Nature Communications dataset, and the Apnea Positive Pressure Long Term Efficacy Study are available from the National Sleep Research Resource at https://sleepdata.org/. All datasets require project approval, prior to use.
MESA and MrOS outcome data are available from https://mesa-nhlbi.org/ and https://mrosonline.ucsf.edu/, after project approval.
Mount Sinai polysomnography data is not available for use.
Model weights can be made available via reasonable request to the authors. Code is available at https://github.com/benmfox/SleepJEPA/.

## 5 Acknowledgments

This work was partially supported by the National Heart, Lung, and Blood Institute of the National Institutes of Health under award number R01HL175992 and the National Center for Advancing Translational Sciences of the National Institutes of Health under award number TL1TR004420. Further, this work was supported in part through the computational and data resources and staff expertise provided by Scientific Computing and Data at the Icahn School of Medicine at Mount Sinai. Research reported in this publication was also supported by the Office of Research Infrastructure of the National Institutes of Health under award number S10OD026880 and S10OD030463. The content is solely the responsibility of the authors and does not necessarily represent the official views of the National Institutes of Health.

The Sleep Heart Health Study (SHHS) was supported by National Heart, Lung, and Blood Institute cooperative agreements U01HL53916 (University of California, Davis), U01HL53931 (New York University), U01HL53934 (University of Minnesota), U01HL53937 and U01HL64360 (Johns Hopkins University), U01HL53938 (University of Arizona), U01HL53940 (University of Washington), U01HL53941 (Boston University), and U01HL63463 (Case Western Reserve University).

The Apnea Positive Pressure Long-term Efficacy Study (APPLES) was supported by the National Heart, Lung, and Blood Institute (U01HL68060).

The Multi-Ethnic Study of Atherosclerosis (MESA) was supported by contracts 75N92025D00022, 75N92020D00001, HHSN268201500003I, N01-HC-95159, 75N92025D00026, 75N92020D00005, N01-HC-95160, 75N92020D00002, N01-HC-95161, 75N92025D00024, 75N92020D00003, N01-HC-95162, 75N92025D00027, 75N92020D00006, N01-HC-95163, 75N92025D00025, 75N92020D00004, N01-HC-95164, 75N92025D00028, 75N92020D00007, N01-HC- 95165, N01-HC-95166, N01-HC-95167, N01-HC-95168 and N01-HC-95169 from the National Heart, Lung, and Blood Institute (NHLBI), and by grants UL1-TR-000040, UL1-TR-001079, and UL1-TR-001420 from the National Center for Advancing Translational Sciences (NCATS). Some mortality data were provided by the Bureau of Vital Statistics, New York City Department of Health and Mental Hygiene. The MESA Sleep Ancillary study was funded by RO1 HL098433 from NHLBI, “Association of Sleep Disorders with Cardiovascular Health Across Ethnic Groups”.

The Wisconsin Sleep Cohort (WSC) Study was supported by the U.S. National Institutes of Health, National Heart, Lung, and Blood Institute (R01HL62252), National Institute on Aging (R01AG036838, R01AG058680), and the National Center for Research Resources (1UL1RR025011).

The National Heart, Lung, and Blood Institute provided funding for the ancillary MrOS Sleep Study, “Outcomes of Sleep Disorders in Older Men,” under the following grant numbers: R01 HL071194, R01 HL070848, R01 HL070847, R01 HL070842, R01 HL070841, R01 HL070837, R01 HL070838, and R01 HL070839.

The Mignot Nature Communications (MNC) research was mostly supported by a grant from Jazz Pharmaceuticals to E.M. Additional funding came from: NIH grant R01HL62252 (to P.E.P.); Ministry of Science and Technology 2015CB856405 and National Foundation of Science of China 81420108002,81670087 (to F.H.); H. Lundbeck A/S, Lundbeck Foundation, Technical University of Denmark and Center for Healthy Aging, University of Copenhagen (to P.J. and H.B.D.S). Additional support was provided by the Klarman Family, Otto Mønsted, Stibo, Vera Carl Johan Michaelsens, Knud Højgaards, Reinholdt W. Jorck and Hustrus and Augustinus Foundations (to A.N.O.).

The National Sleep Research Resource was supported by the National Heart, Lung, and Blood Institute (R24 HL114473, 75N92019R002).

The Human Sleep Project has received support from the Glenn Foundation and the American Federation of Aging Research (AFAR) through the 2018 Glenn / AFAR Award for Medical Research Breakthroughs in Gerontology (BIG) (2018), the American Academy of Sleep Medicine (AASM) through a 2019 Strategic Research Award, the National Institutes of Health (NIH) (R01NS102190, R01NS102574, R01NS107291, RF1AG064312, RF1NS120947, R01AG073410, R01HL161253, R01NS126282, R01AG073598), the National Science Foundation (NSF 2014431), and through the Henry and Allison McCance Center for Brain Health.

## A Extended Data

**Table 1:**
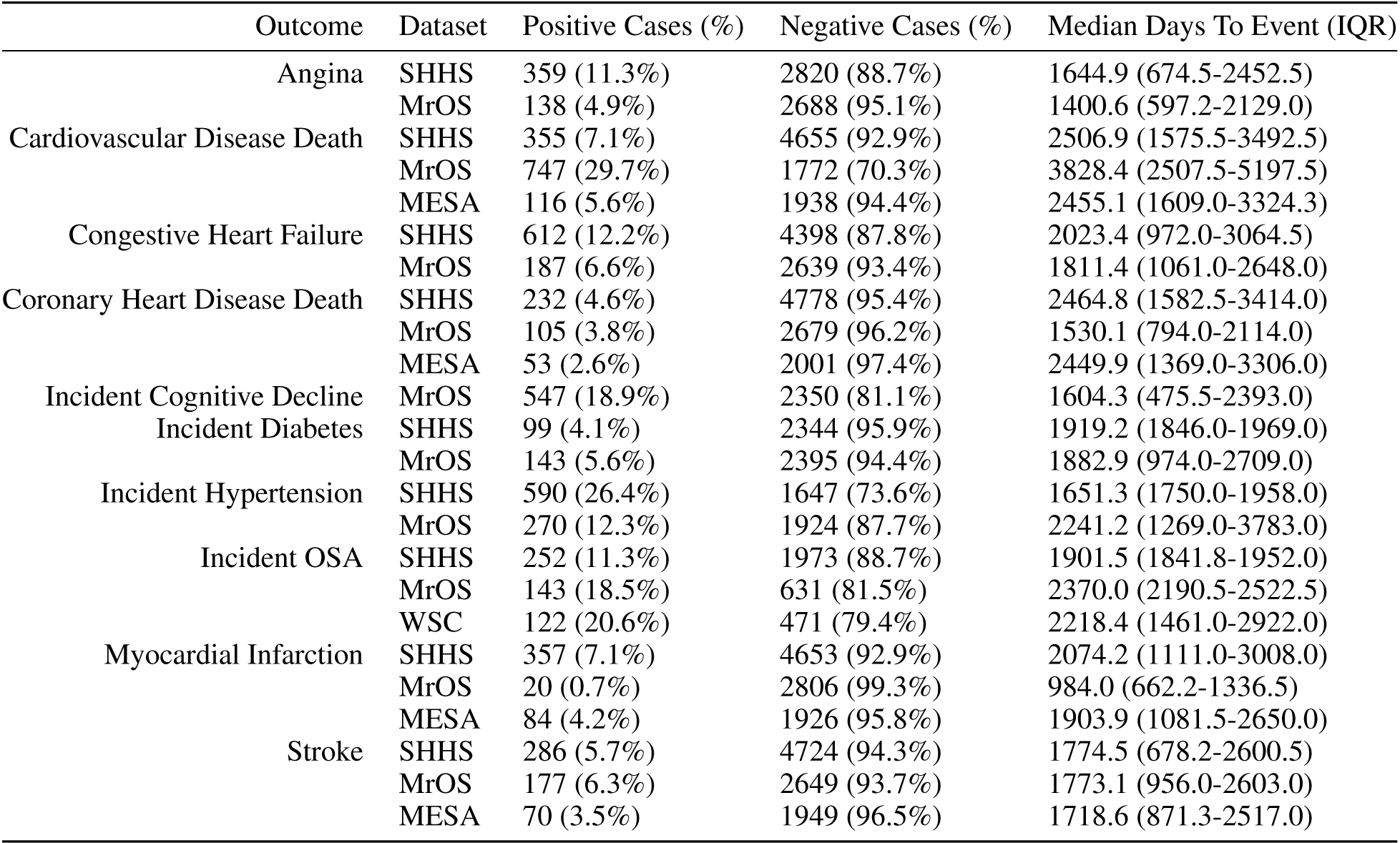
Long-term disease risk distributions for finetuning. Outcomes were provided by the study cohort datasets. “Incident” outcomes were calculated manually from follow up visits, since the baseline sleep study. Training, validation, and held-out testing followed identical splitting ratios for all outcomes: 72%, 8%, 20%, respectively.

**Figure 1:**
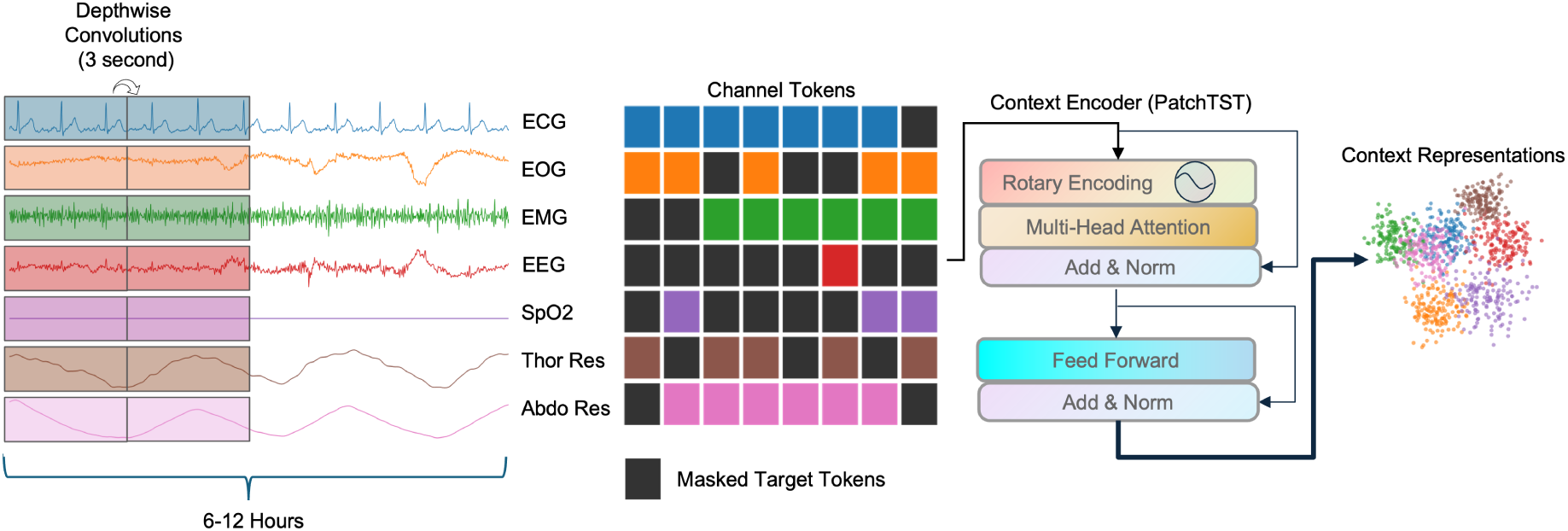
SleepJEPA Context Encoder. During JEPA training, tokenized patches of signal data are randomly selected as targets for prediction. Targets are masked and only context tokens are passed through the context encoder to generate context representations.

**Figure 2:**
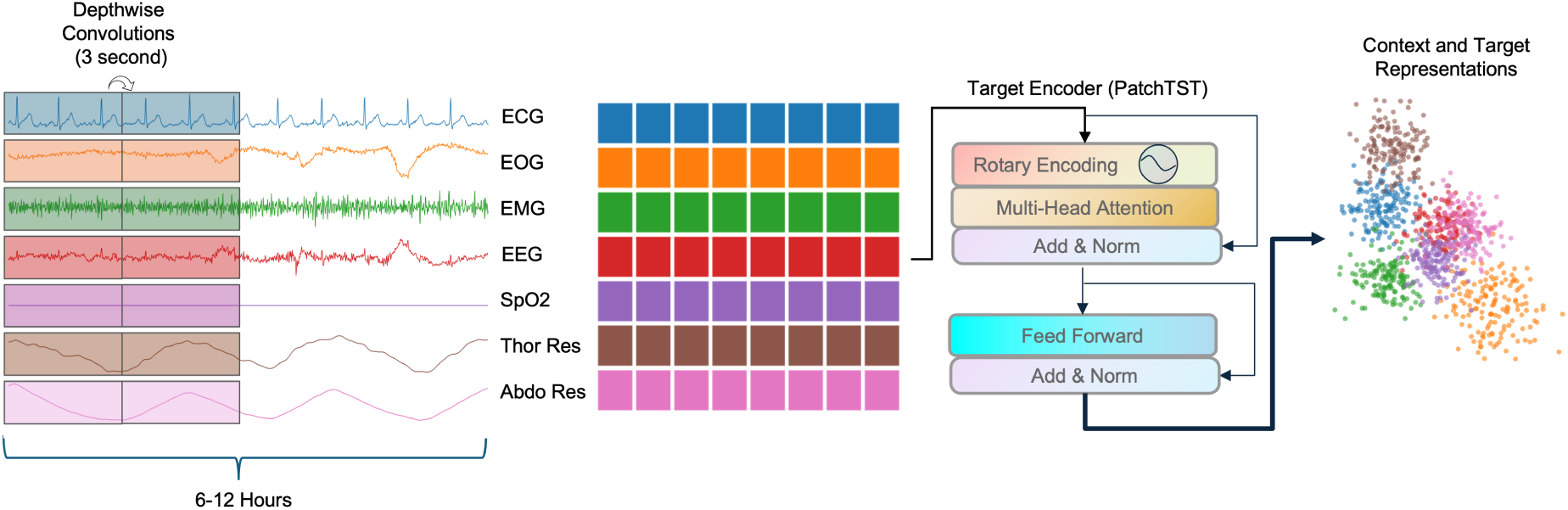
SleepJEPA Target Encoder. During JEPA training, the target encoder receives the full input without masking, creating context and target representations. Note that the target encoder is identical to the context encoder, but its weights lag the context encoder weights and are updated by exponentially weighted moving average after each batch.

**Figure 3:**
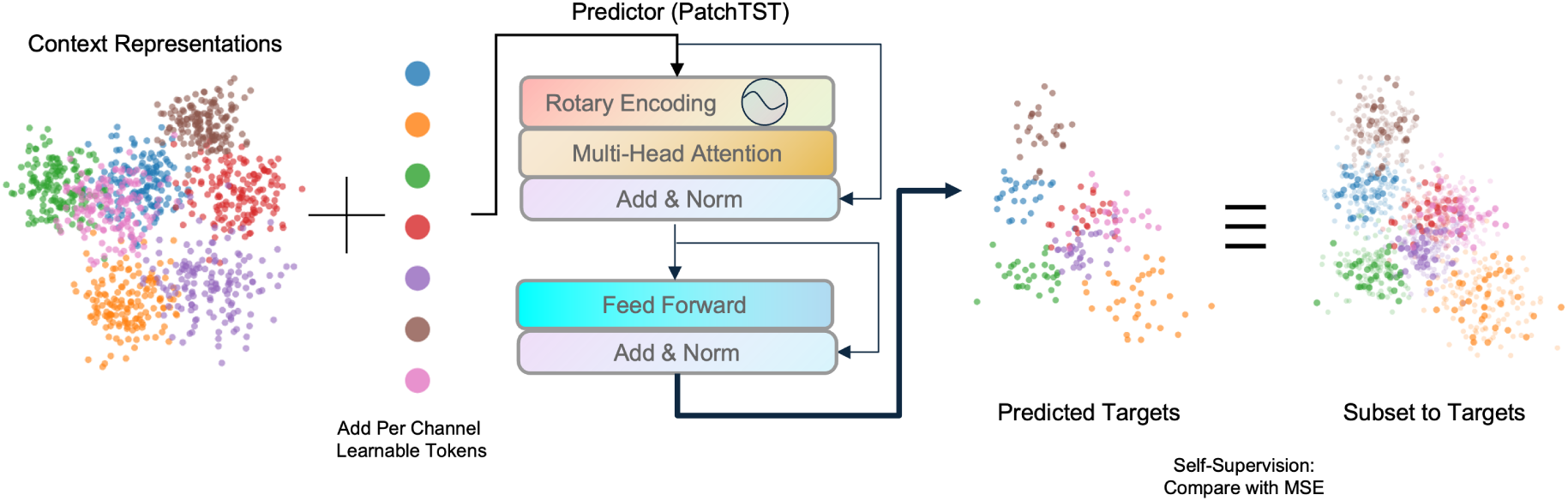
SleepJEPA Predictor. During JEPA training, the context representations are combined with per channel, learnable mask tokens and then passed through a predictor network (a smaller network, compared to the context and target encoders). The predictions are then compared to the target encoders output, after subsetting the outputs to the targets. The two are compared via an MSE loss function.

**Figure 4:**
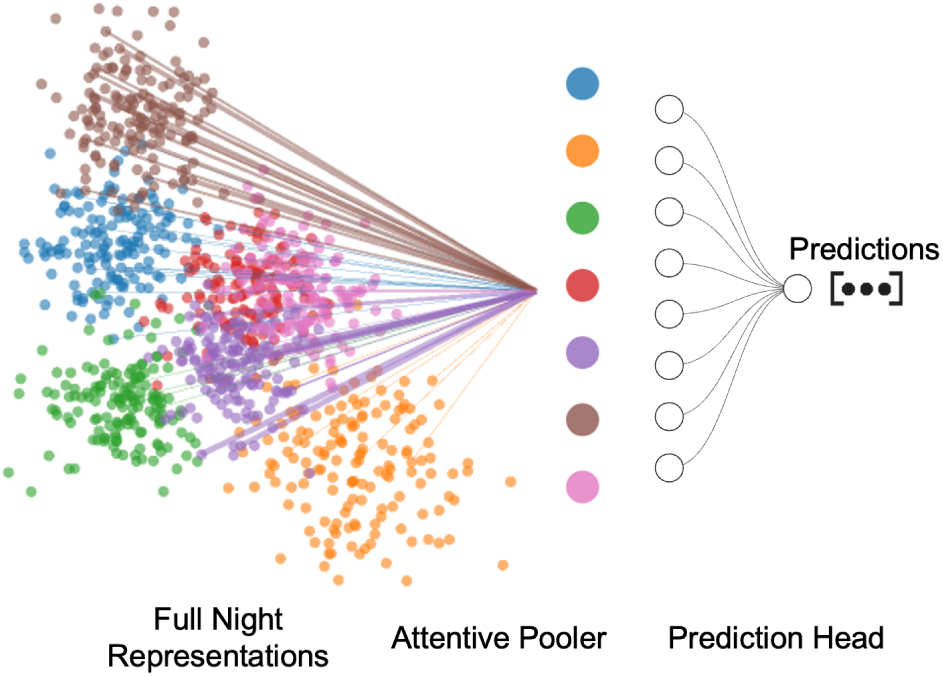
Attentive Finetuning Head. The attentive head pools patches across all channels with a full cross attention block into a single vector per channel. Following, the channel vectors are passed through a final prediction head, which is task dependent.

**Figure 5:**
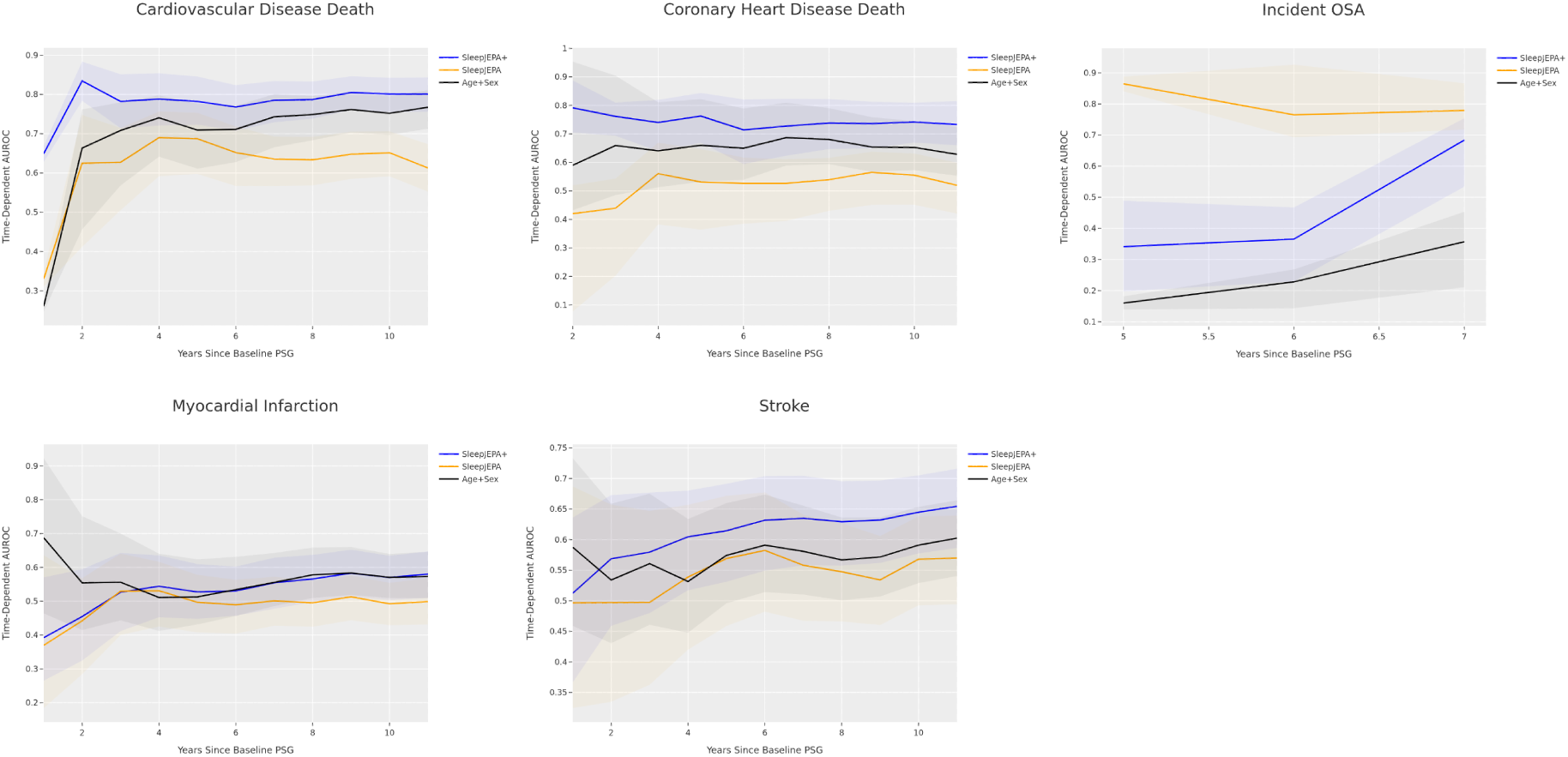
C/D AUC plots for independent test sets for available outcomes, up to 15 years. SleepJEPA+, SleepJEPA, and the baseline demographics models are reported. All models used MESA as an independent test set, except for incident OSA, which used WSC. 95% bootstrapped confidence intervals (n = 999 resamples) are shaded for each model.

**Figure 6:**
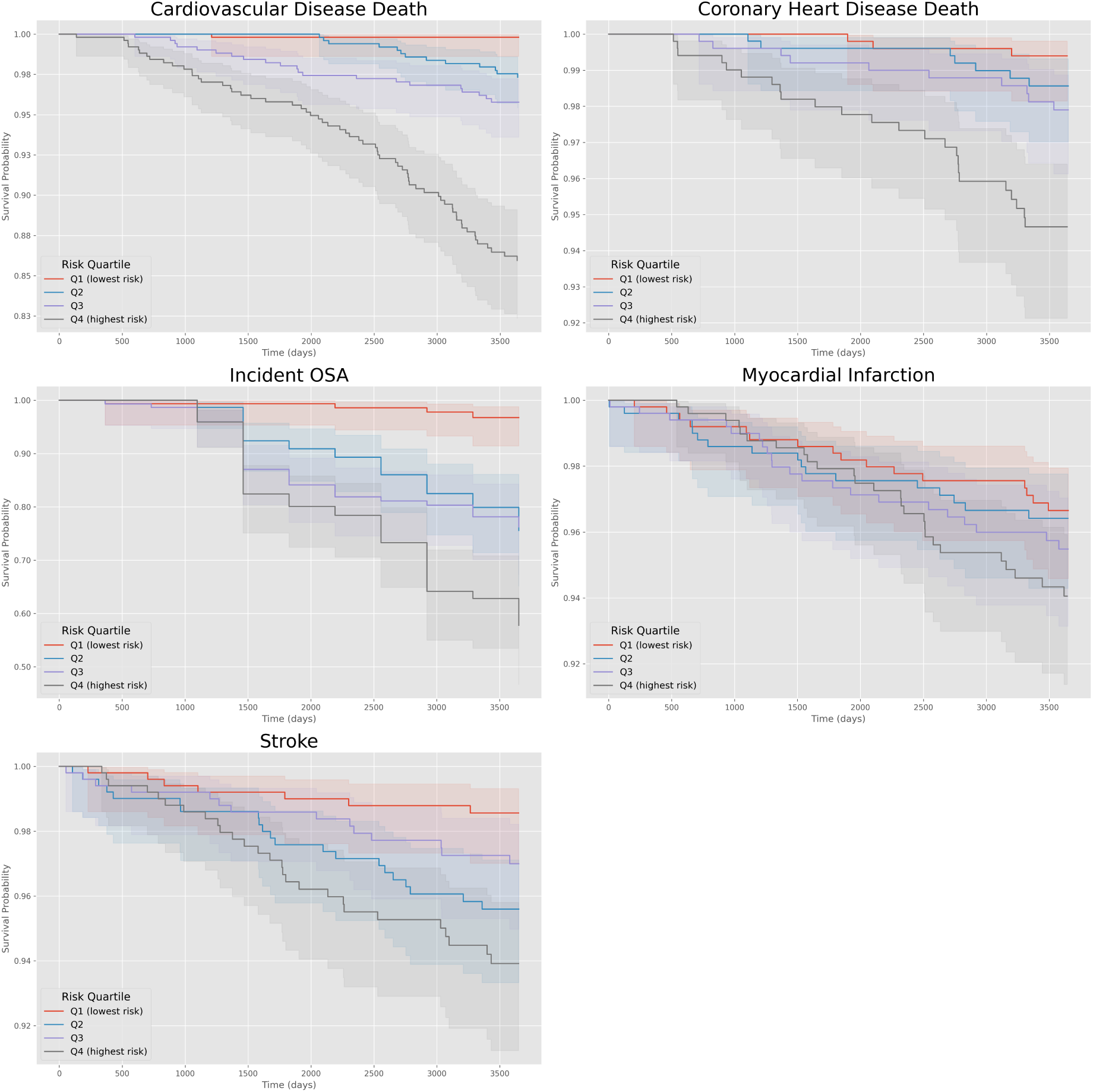
Survival curves at 10 years for all outcomes examining SleepJEPA+ risk quartiles on the SHHS held-out test set and MrOS held-out test set. Probabilities were calibrated using Platt scaling per model-outcome using the validation set prior to survival monitoring. 95% confidence intervals are displayed in bands.

