## Supplemental Material for "SleepJEPA: Learning the latent world of sleep with at-home sleep data to estimate disease risk"

SUPPLEMENTAL MATERIALS

#### 1 Data and Code Sources

We utilize the Human Sleep Project (HSP) (Westover et al.), the Mount Sinai Polysomnography Database (MS-PSG), the Sleep Heart Health Study (SHHS) (Quan et al., 1997; Zhang et al., 2018), the Wisconsin Sleep Cohort (WSC) (Young, 2009), the Osteoporotic Fractures in Men Study (MrOS) (Blackwell et al., 2011), the Multi-Ethnic Study of Atherosclerosis (MESA) (Chen et al., 2015; Zhang et al., 2018), the Mignot Nature Communications (MNC) dataset (Stephansen et al., 2018; Zhang et al., 2018), and the Apnea Positive Pressure Long Term Efficacy Study (APPLES) (Quan et al.). All studies used deidentified, retrospective PSG data made available through the National Sleep Research Resource (NSRR) at `https://sleepdata.org/` (Zhang et al., 2018), the Brain Data Science Platform at `https://bdsp.io/content/hsp/2.0/` (Westover et al.), or Mount Sinai's Sleep, Circadian, and Analysis Group. Outcome data was acquired for MESA from the MESA coordinating center and MrOS outcome data was acquired at `https://mrosonline.ucsf.edu/`. Data access was approved for use. Model code can be found at `https://github.com/benmfox/SleepJEPA/` and weights can be made available upon reasonable request to the authors.

#### 2 Methods

##### 2.1 Sleep Study Data Preparation

SHHS, MESA, and MrOS were Type II, unattended at-home sleep studies. HSP, MS-PSG, WSC, MNC, and APPLES were Type I research or clinic sleep studies. EDF files were read and at-home sleep signal channels were extracted and stored in zarr format (Miles et al., 2024). A single electroencephalogram (EEG, C4-M1 or C3-M2), left electrooculogram (EOG), chin electromyogram (EMG), augmented lead II electrocardiogram (ECG), oxygen saturation (SpO2), thorax respiratory inductance plethysmography (RIP), and abdominal RIP were extracted from all studies. Sleep studies shorter than 6 hours or longer than 12 hours were excluded from analysis. Additionally, if a single extracted channel in a sleep study was greater than or equal to 50% constant or null values, it was removed from the pretraining dataset. All channels were resampled to 128 Hz. Signals were processed using infinite impulse response (IIR) elliptic filters with an order of 16, max passband of 1 dB, and minimum stopband of 40 dB to remove noise and artifacts, as previously described (Brink-Kjaer et al., 2022). EEG and EOG channels had a bandpass filter of 0.3-45 Hz, EMG a high pass filter of 10 Hz, ECG a high pass filter of 0.3 Hz, and thorax and abdominal RIP channels a bandpass filter of 0.1-15 Hz. SpO2 had no filter applied. Following filtering, percentile normalization was performed, scaling every channel to -1 (5th percentile) to 1 (95th percentile). SpO2 was scaled from -1 (60% saturation) to 1 (100% saturation). After preprocessing the input size per-channel length to SleepJEPA ranged from 2,764,800 (6 hours) to 5,529,600 (12 hours). To account for varying length sleep studies in SleepJEPA, PyTorch's nested tensor module was utilized, avoiding unnecessary padding of sleep signals (Paszke et al., 2019).

##### 2.2 SleepJEPA Pretraining

SleepJEPA was pretrained with HSP, MS-PSG, WSC, MNC, and APPLES datasets. The initial learning rate was set to 1.6e-5, and the learning rate was adjusted according to a one cycle scheduler with a max learning rate of 0.0004, 20% warmup period, and a cosine annealing reduction to 1.6e-7. Momentum was not cycled. AdamW was used for optimization with a weight decay scheduler starting at 0.04 and ending at 0.4 over the course of training, as performed in the original JEPA implementation. The target encoder weights were updated with an exponentially weighted moving average of the context encoder weights, starting at 0.998 and reaching 1 by the end of training. Gradients were clipped to a global norm to be less than or equal to 1, to avoid exploding gradients. Augmentations were applied to the input data randomly (p=0.5) during pretraining. Sleep studies were randomly cropped and timepoints were randomly masked or corrupted with noise to encourage generalization across variable-length recordings and a robustness to noise. SleepJEPA pretraining was performed across 3 nodes, with 4 NVIDIA H100nvl GPUs per node, and a batch size of 2 (effective batch size of 24).

##### 2.3 SleepJEPA Architecture

After signal preprocessing, signals are tokenized into 3 second patches via a single per-channel convolution, which creates "tokens" of sleep signals for input into the transformer architecture. We chose 3 second tokens because of previous work showing sleep states and features can be derived from shorter segments of sleep (Lechat et al., 2022; Younes et al., 2015). Target tokens are selected at random per sample-channel (20%-80% masking), and the remaining tokens are used as context. While 20-80% is a broad range, recent works suggest that exposing models to varying masking ratios is optimal for learning robust representations (Dong et al., 2025). Context tokens are passed through an adapted or signal data patch time series transformer (PatchTST) (Fox et al., 2025; Nie et al., 2023) comprising 8 layers and 8 heads, with a feature dimension of size 768 to embed the signals. Notably, channels are embedded independently, leaving it to the attention mechanism to learn cross-channel relationships and allow for variable channel input. Rotary positional embeddings are applied to learn relative and absolute positional encodings across patches (Su et al., 2024). Per-channel mask tokens are appended to context encodings to encourage channel independent feature learning. A predictor PatchTST network of 4 layers, 8 heads, and dimension of 384 estimates the targets. The true target tokens and predicted targets are compared with a mean squared error loss function. An overview of the context encoder, target encoder, and predictor are detailed in Extended Data Figures 1, 2, and 3. In total, the SleepJEPA context encoder and predictor contained 66,464,256 parameters. The context encoder was used for downstream representation generation and finetuning and contains 58,772,736 parameters. The input size to the context encoder is 7 channels x [6-12] hours of signal data. The context encoder outputs a tensor of shape 7 channels x n patches (3 sec) x 768. The number of patches varies per sample and is handled by the nested tensor module in PyTorch.

##### 2.4 Sleep Staging Data Preparation

Sleep stages were trained and evaluated on SHHS, MrOS Visit 1, and WSC, and independently tested on MESA, MrOS Visit 2, and APPLES. Sleep stages across all studies were labeled using Rechtshaffen and Kales criteria (Wolpert, 1969) by a randomly assigned single scorer in a central reading center. Hypnograms indicating REM and NREM (wake, N1, N2, N3) were extracted and time matched to sleep signal data, with 30 second labels. 30 second segments labeled as movement or unknown were ignored during training. Stage 4 sleep was truncated into stage 3. Only sleep studies with at least one segment of wake, stage 2, and REM were utilized during training, validation, and testing.

##### 2.5 Sleep Stage Classification Architecture and Training Details

A GRU classifier was trained using frozen SleepJEPA representations to estimate sleep stages with a cross-entropy loss function. A 2-layer, bidirectional GRU with a hidden size of 768, and dropout of 0.1 followed by a multi-layer perceptron (MLP) was used to estimate sleep stages. While the GRU modeled the relationship between 3 second patches and predicted sleep stages per 3 second patch, patch predictions were averaged across 30 seconds (every 10 patches) for comparison to target labels. The sleep stage classification model had 40,132,627 trainable parameters. The initial learning rate was set to 4e-5, and the learning rate was adjusted according to a one cycle scheduler with a maximum learning rate of 0.001, 30% warmup period, and a cosine annealing reduction to 4e-9. Momentum was cycled. AdamW was used for optimization and weight decay was set to 0.0001. Gradients were clipped to a global norm to be less than or equal to 1, to avoid exploding gradients. The batch size was set to 10, and the model was trained for 25 epochs on a single NVIDIA H100nvl GPU. Validation AUC was monitored during training at the end of each epoch, and the epoch-model with the highest validation AUC was chosen for testing.

##### 2.6 Daytime Sleepiness and Narcolepsy Outcomes

Objective daytime sleepiness was derived from MSLT in the WSC dataset, where the mean sleep latency was less than or equal to 8 minutes. Narcolepsy outcomes were provided by the MNC study authors and have type 1 narcolepsy diagnoses. Narcolepsy labels reflect mean sleep latency of less than or equal to 8 minutes and presence of at least 2 sleep onset REM periods (REM latency of less than or equal to 15 minutes in at least 2 naps). Given the similarities between the two sleepiness outcomes, MNC was used as an independent test set for objective daytime sleepiness, and WSC was used as an independent test set for narcolepsy.

##### 2.7 Age Regression and Daytime Sleepiness Classification Architectures and Training Details

Age estimation was trained with SHHS visits 1 and 2 and MrOS visit 1 and independently evaluated on WSC and MESA. For both age and daytime sleepiness finetuning models, an attentive pooler including a full cross attention block followed by a linear layer was used for training. For age, the loss function was mean squared error, and for daytime sleepiness outcomes, the loss function was class-weighted binary cross entropy. Each model had 7,094,017 trainable parameters. The initial learning rate was set to 1e-3 and followed a cosine annealing with warm restarts learning rate scheduler, with 5 epochs until the first restart, and a multiplier of 2. The minimum learning rate was set to 1e-6. Again, AdamW was used for optimization and weight decay was set to 0.001. Gradients were clipped to a global norm of less than or equal to 1. The batch size was set to 16, and each model was trained for 25 epochs on a single NVIDIA H100nvl GPU. For age, the validation MSE was monitored during training at the end of each epoch, and the epoch-model with the lowest validation MSE was chosen for testing. For daytime sleepiness outcomes, validation AUC was monitored during training at the end of each epoch, and the epoch-model with the highest validation AUC was chosen for testing.

##### 2.8 Long-term Disease Risk Estimation

SHHS, MrOS, and MESA provided adjudicated outcomes and time to event for angina, cardiovascular (CV) disease death, congestive heart failure, coronary heart disease death, myocardial infarction, and stroke.

For details on SHHS event labeling, see the protocol at `https://sleepdata.org/datasets/shhs/files/m/browser/documentation/SHHS_CVD_Outcomes_Protocol.pdf`.

For details on MESA event labeling, see `https://www.ncbi.nlm.nih.gov/projects/gap/cgi-bin/GetPdf.cgi?id=phd001674.1`.

For details on MrOS event labeling, see `https://mrosonline.ucsf.edu/DataRelease/ReleasedDatasets`.

Incident outcomes of hypertension, OSA, diabetes, and cognitive impairment were derived from follow-up information after the baseline study. For all incident outcomes, subjects were required to be a "negative" case at the baseline study, given the following criteria for each outcome. For incident hypertension, if the subject had a blood pressure medication change and self-reported being treated by a doctor and had a blood pressure reading higher than 140/90 at the follow-up visit, they were marked as a positive case. Again, the date of the follow up visit was used as the time to event for positive cases, and the date of last verified negative case was used as the censoring date for negative cases. For incident diabetes, if the subject began taking oral hypoglycemic agents or insulin and indicated they were being treated by a doctor (MrOS only), they were marked as a positive case. For incident OSA, if the subject indicated they are being treated for sleep apnea by a doctor (MrOS and SHHS only) and had an apnea hypopnea index >= 30 at a follow up visit, they were marked as a positive case. Finally, for incidental cognitive decline, a change in the Modified Mini-Mental State (3MS) exam (Teng and Chui, 1987) >= 1.5x the mean change from baseline (in the negative direction), self-report of dementia diagnosis by a doctor, or dementia medication use was used to derive outcomes, as previously described (Cavaillès et al., 2025). Those with a baseline 3MS < 80 or dementia medication use were excluded.

##### 2.9 Long-term Disease Risk Architecture and Training Details

For each long-term disease, an attentive pooler including a full cross attention block followed by a self-attention layer and linear classifier was used for training. Each model comprised 14,181,889 trainable parameters. If demographics were included in the model (or for the demographics only baseline), the model utilized a MLP to encode age with 559 parameters (32 hidden size). Age was normalized based on training dataset statistics prior to input. 2 parameters were optimized to encode sex. The attentive classifier outputs N predictions per batch sample, where N is the number of years to estimate risk (times are discretized to years, if given in days). Similarly, the age MLP outputs N year risk estimates. The total hazard was calculated by summing SleepJEPA's representation generated hazards (from the attentive classifier), with the age hazards (from the MLP), and the single parameter sex hazard. Finally, to model the interaction between the representations and time, and age and time, additional N trainable parameters were added to the total hazard. Thus, each prediction represents the estimated hazard at each year, while considering the effect of time. To optimize the model, a discrete hazard loss function is employed:

$$\mathcal{L}_{\text{disc}}(h) = -\sum_{j=1}^{J} \left( \sum_{i=1}^{d_j} \ln\left(h_j^i\right) + \sum_{i=d_j+1}^{r_j} \ln\left(1 - h_j^i\right) \right) \tag{1}$$

Where $J$ is the number of years, $d_j$ is the number of people who experience an event in that year, and $r_j$ is the number of people who are censored in that year. $h$ is the predicted hazard. The loss function helps the model learn to maximize the hazard for those that experience an event sooner across all bins. The loss function was adapted from pycox (`https://github.com/havakv/pycox`).

To train each model, the initial learning rate was set to 1e-3 and followed a cosine annealing with warm restarts learning rate scheduler, with 5 epochs until the first restart, and a multiplier of 2. The minimum learning rate was set to 1e-6. AdamW was used for optimization and weight decay was set to 0.0001. Gradients were clipped to a global norm of less than or equal to 1. The batch size was set to 16 and each model was trained for 10 epochs on a single NVIDIA H100nvl GPU. C-index, IBS, and mean C/D AUC were monitored on the validation set throughout training, and the epoch-model with the highest combined score of the three metrics (C-Index + AUC + (1-IBS)) was chosen.

Two additional models were trained per outcome, following identical training procedures. The baseline demographics model followed an identical architecture as described above; however, the SleepJEPA representation hazards were not calculated and added to the demographic hazards. Consequently, the SleepJEPA representations only model followed an identical architecture; however, the demographic hazards were not calculated and added to the SleepJEPA representation hazards.

##### 2.10 Training Details

All models were trained using PyTorch (`https://pytorch.org/`) and PyTorch Lightning (`https://lightning.ai/`). Weights and Biases (`https://wandb.ai/`) was used for monitoring. Additionally, all models were trained with FlashAttentionV2 and mixed precision.

##### 2.11 Evaluation Details

Models were evaluated with torchmetrics (`https://lightning.ai/docs/torchmetrics/stable/`) and survival metrics were evaluated with scikit-survival (`https://scikit-survival.readthedocs.io/`) and torchsurv (`https://opensource.nibr.com/torchsurv/`).

##### 2.12 Integrated Gradients

The integrated gradients method was implemented with Captum, a model interpretability package for PyTorch (`https://captum.ai/`). Positive samples for each outcome from the SHHS held-out test set or MrOS (for cognitive decline) were identified from the test set with a positive case between 5 and 10 years after the baseline sleep study. The top five samples with the highest SleepJEPA+ estimated risk at 10-years were extracted to analyze attributions. Five age and gender matched negative samples were selected for baselines for the integrated gradients calculation. The frozen SleepJEPA+ representations were used as input to the integrated gradients method with 512 steps. Attribution scores were normalized using the 99th percentile of the highest attribution across all batch attributions. Attributions were plotted on a heatmap to display aggregated attributions by sleep stage and total aggregations per channel.

#### 3 Supplementary Figures

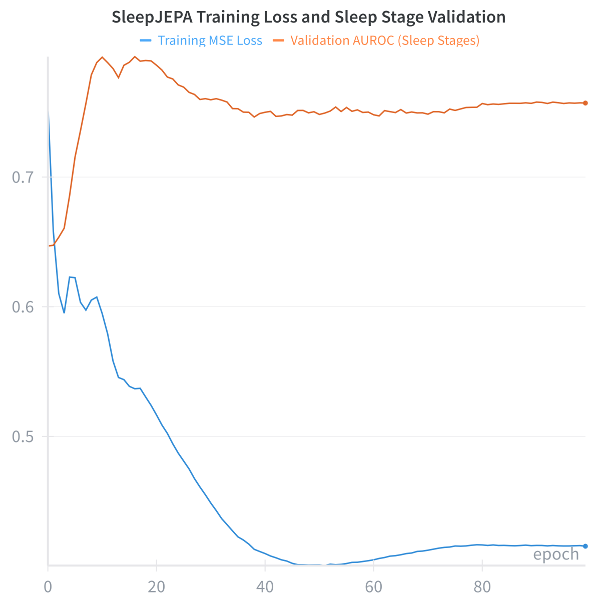


Supplementary Figure 1: SleepJEPA Training Loss and Sleep Stage Validation. SleepJEPA training was monitored by MSE training loss and sleep stage classification per epoch with a validation set. The epoch-model with the highest AUC on sleep stage prediction was used for finetuning all downstream tasks. This occurred at epoch 16.

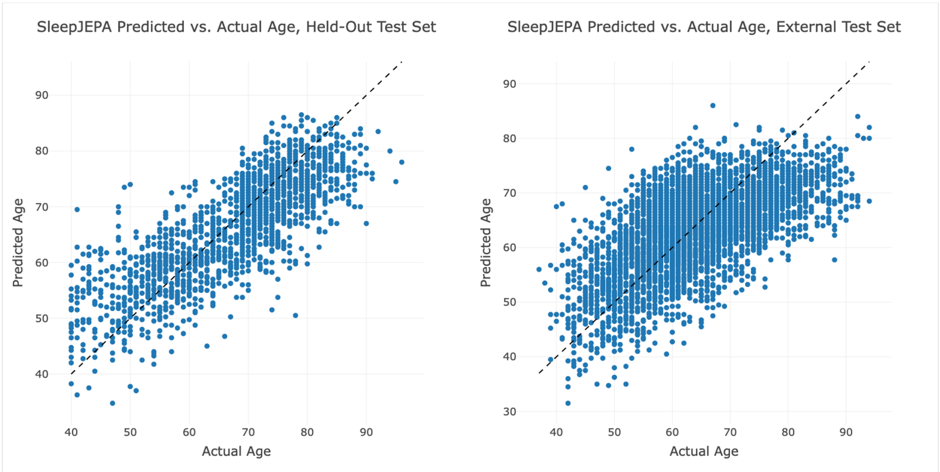


Supplementary Figure 2: SleepJEPA Predicted Age Vs Actual Age Plots. Predicted and actual age are shown for the held-out test set (left) and independent test set (right).

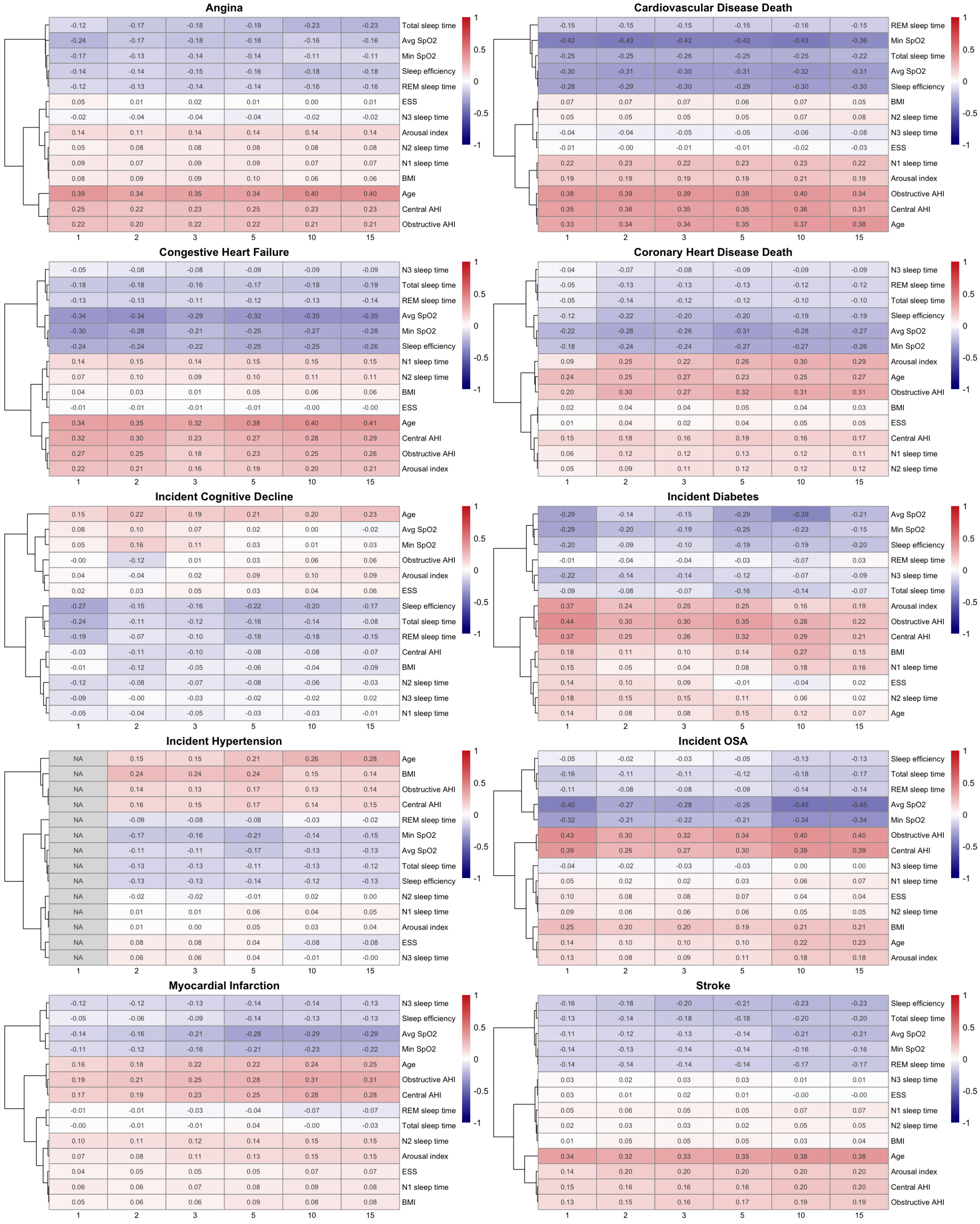


Supplementary Figure 3: SleepJEPA representations predicted cumulative risk correlations at different time horizons, up to 15 years. Cumulative risk scores' Pearson correlation was calculated with normalized demographic and baseline PSG derived features. Hierarchical clustering was performed using the ward method across rows. Correlations are shown across different forecasting horizons for the SHHS held-out test set and MrOS held-out test set (for cognitive decline).

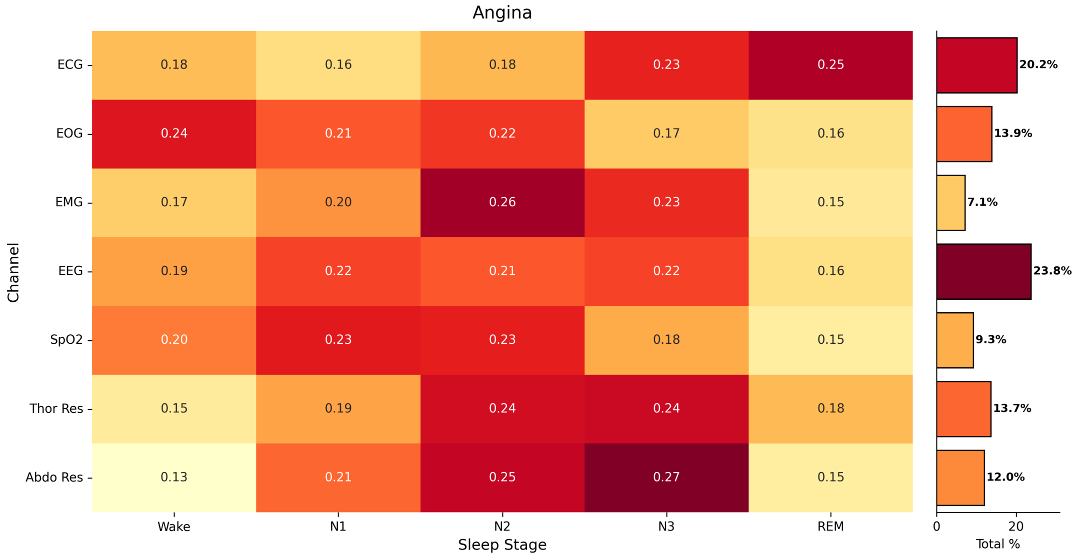


Supplementary Figure 4: SleepJEPA+ representation attributions for angina classification on 5 positive event sample from the SHHS held-out test set with high predicted risk by sleep stage. The events occurred 5-10 years after the baseline sleep study. Attribution scores are normalized across all samples and channels with the 99th percentile largest attribution value. The percentages of aggregated channel attributions are displayed in the bar chart to the right of each channel.

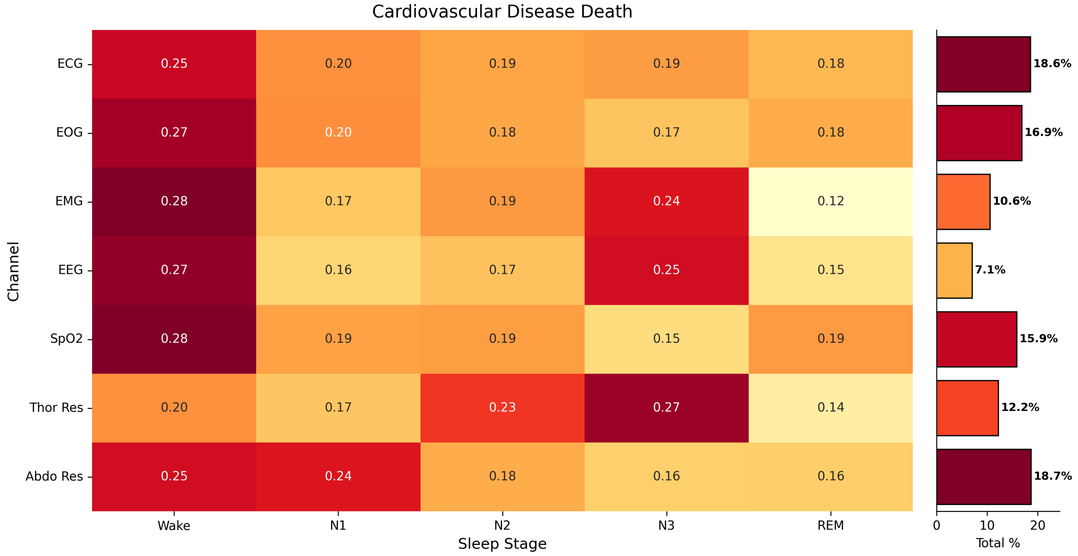


Supplementary Figure 5: SleepJEPA+ representation attributions for CV disease death classification on 5 positive event sample from the SHHS held-out test set with high predicted risk by sleep stage. The events occurred 5-10 years after the baseline sleep study. Attribution scores are normalized across all samples and channels with the 99th percentile largest attribution value. The percentages of aggregated channel attributions are displayed in the bar chart to the right of each channel.

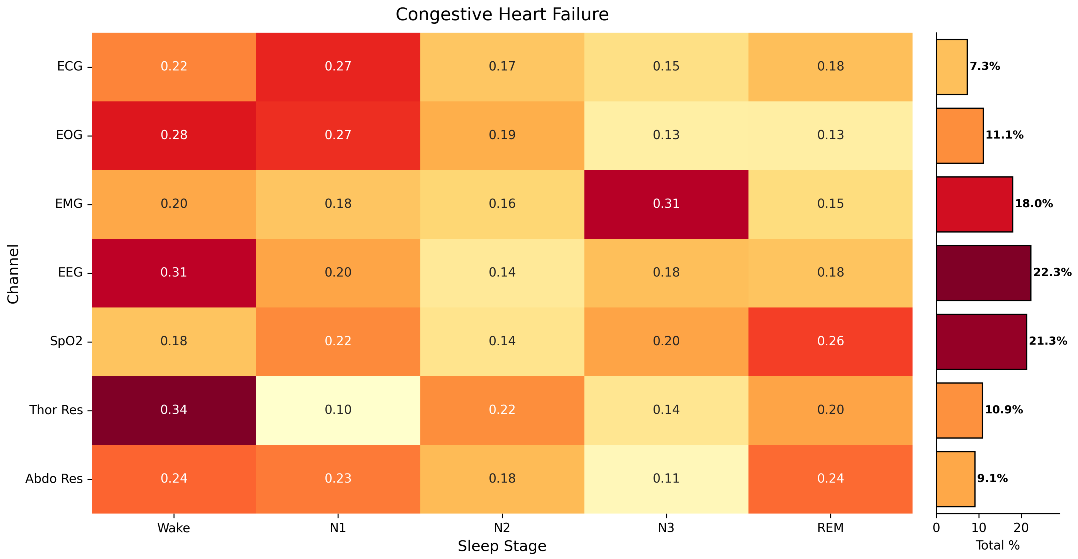


Supplementary Figure 6: SleepJEPA+ representation attributions for congestive heart failure classification on 5 positive event sample from the SHHS held-out test set with high predicted risk by sleep stage. The events occurred 5-10 years after the baseline sleep study. Attribution scores are normalized across all samples and channels with the 99th percentile largest attribution value. The percentages of aggregated channel attributions are displayed in the bar chart to the right of each channel.

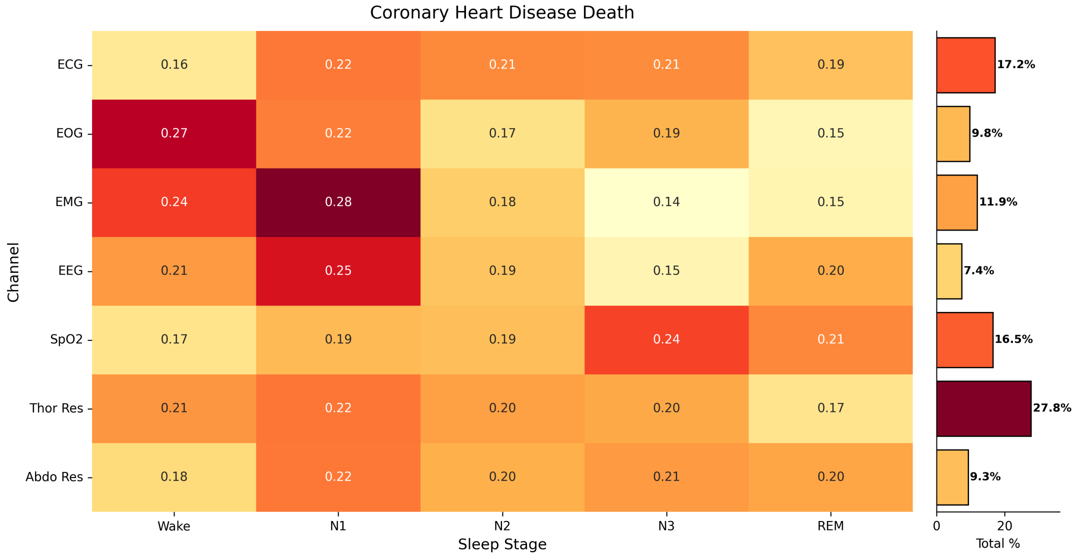


Supplementary Figure 7: SleepJEPA+ representation attributions for coronary heart disease death classification on 5 positive event sample from the SHHS held-out test set with high predicted risk by sleep stage. The events occurred 5-10 years after the baseline sleep study. Attribution scores are normalized across all samples and channels with the 99th percentile largest attribution value. The percentages of aggregated channel attributions are displayed in the bar chart to the right of each channel.

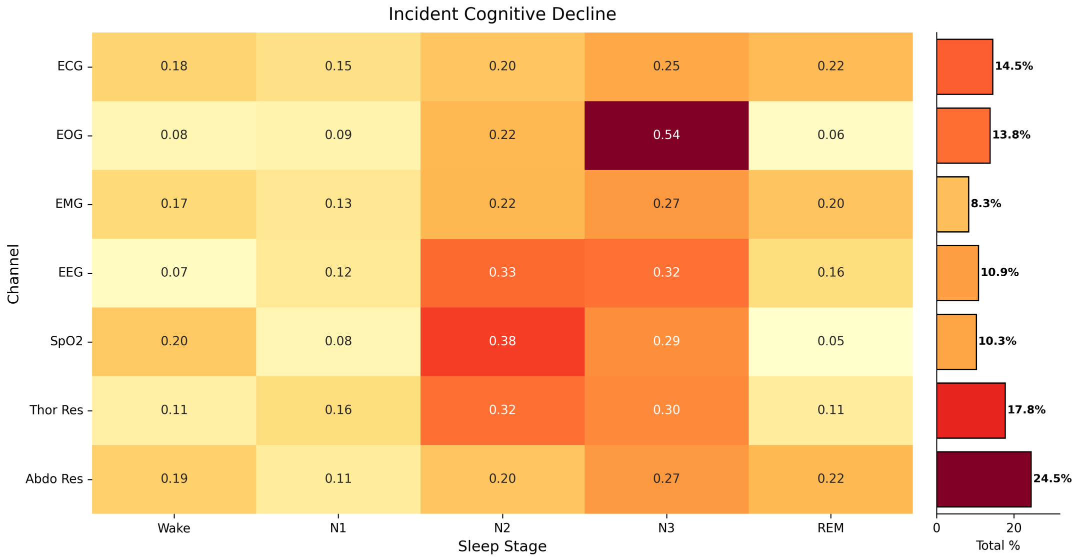


Supplementary Figure 8: SleepJEPA+ representation attributions for incident cognitive decline classification on 5 positive event sample from the MrOS held-out test set with high predicted risk by sleep stage. The events occurred 5-10 years after the baseline sleep study. Attribution scores are normalized across all samples and channels with the 99th percentile largest attribution value. The percentages of aggregated channel attributions are displayed in the bar chart to the right of each channel.

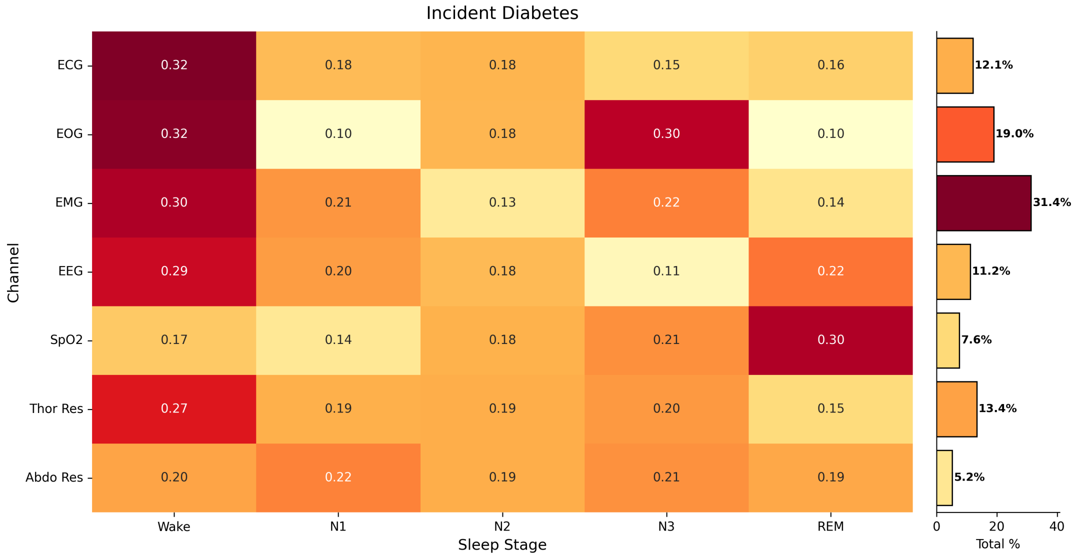


Supplementary Figure 9: SleepJEPA+ representation attributions for incident diabetes classification on 5 positive event sample from the SHHS held-out test set with high predicted risk by sleep stage. The events occurred 5-10 years after the baseline sleep study. Attribution scores are normalized across all samples and channels with the 99th percentile largest attribution value. The percentages of aggregated channel attributions are displayed in the bar chart to the right of each channel.

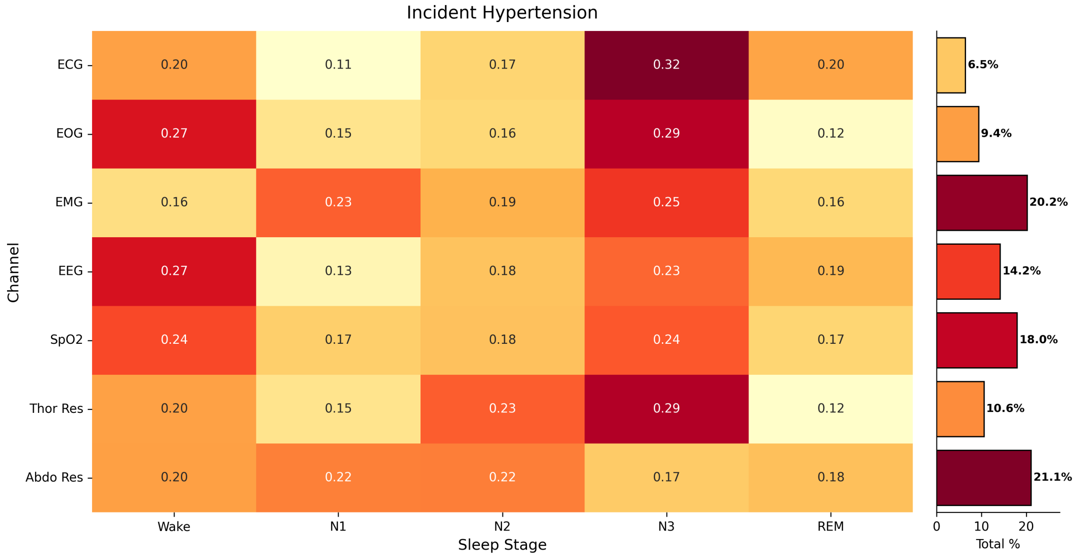


Supplementary Figure 10: SleepJEPA+ representation attributions for incident hypertension classification on 5 positive event sample from the SHHS held-out test set with high predicted risk by sleep stage. The events occurred 5-10 years after the baseline sleep study. Attribution scores are normalized across all samples and channels with the 99th percentile largest attribution value. The percentages of aggregated channel attributions are displayed in the bar chart to the right of each channel.

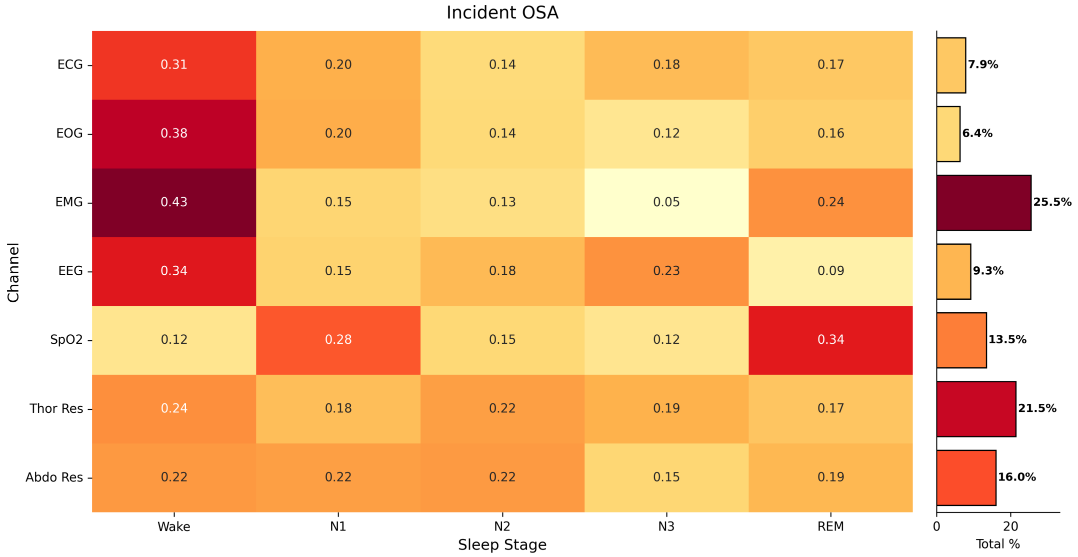


Supplementary Figure 11: SleepJEPA+ representation attributions for incident OSA classification on 5 positive event sample from the SHHS held-out test set with high predicted risk by sleep stage. The events occurred 5-10 years after the baseline sleep study. Attribution scores are normalized across all samples and channels with the 99th percentile largest attribution value. The percentages of aggregated channel attributions are displayed in the bar chart to the right of each channel.

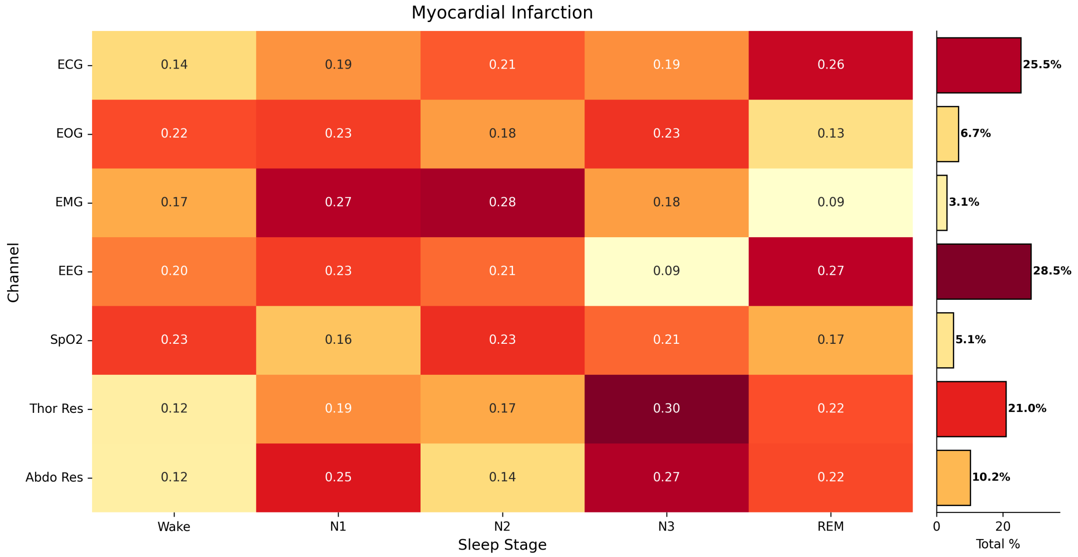


Supplementary Figure 12: SleepJEPA+ representation attributions for myocardial infarction classification on 5 positive event sample from the SHHS held-out test set with high predicted risk by sleep stage. The events occurred 5-10 years after the baseline sleep study. Attribution scores are normalized across all samples and channels with the 99th percentile largest attribution value. The percentages of aggregated channel attributions are displayed in the bar chart to the right of each channel.

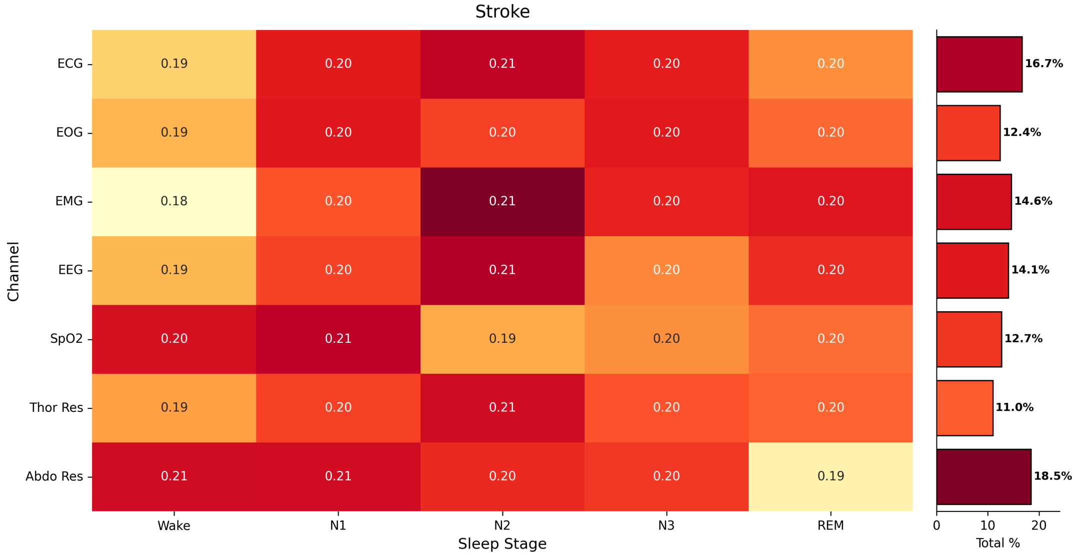


Supplementary Figure 13: SleepJEPA+ representation attributions for stroke classification on 5 positive event sample from the SHHS held-out test set with high predicted risk by sleep stage. The events occurred 5-10 years after the baseline sleep study. Attribution scores are normalized across all samples and channels with the 99th percentile largest attribution value. The percentages of aggregated channel attributions are displayed in the bar chart to the right of each channel.

### 4 Supplementary Tables

| Dataset | n Studies | Wake | N1 | N2 | N# | REM | Split |
|---|---|---|---|---|---|---|---|
| SHHS Visit 1 | 5575 | 28.90% | 3.72% | 40.84% | 12.61% | 13.94% | Training/Validation/Test |
| MrOS Visit 1 | 2845 | 45.70% | 3.65% | 34.08% | 6.08% | 10.49% | Training/Validation/Test |
| SHHS Visit 2 | 2608 | 37.86% | 3.49% | 35.95% | 9.82% | 12.88% | Training/Validation/Test |
| MESA | 2010 | 43.28% | 7.84% | 32.84% | 5.75% | 10.30% | Independent Test |
| WSC | 1789 | 18.47% | 8.33% | 53.74% | 5.78% | 13.68% | Training/Validation/Test |
| APPLES | 1044 | 24.21% | 13.99% | 46.02% | 2.34% | 13.44% | Independent Test |
| MrOS Visit 2 | 996 | 56.38% | 5.17% | 27.23% | 2.91% | 8.30% | Independent Test |

Supplementary Table 1: Sleep staging distributions. Training, validation, and held-out testing were split across all cohorts at 72%, 8%, and 20%, respectively. Labeled hypnograms were required to have at least one stage of wake, N2, and REM to be included in training, validation, and testing.

| Dataset | Outcomes | Positive Cases (%) | Negative Cases (%) |
|---|---|---|---|
| WSC | Objective Daytime Sleepiness | 380 (26.3%) | 1066 (73.7%) |
| MNC | Type 1 Narcolepsy | 44 (8.8%) | 454 (91.2%) |

Supplementary Table 2: Daytime Sleepiness outcome distributions. Objective daytime sleepiness was derived from MSLT, when the mean sleep latency was less than or equal to 8 minutes. Narcolepsy labels were provided by the MNC study and reflect mean sleep latency of less than or equal to 8 minutes and presence of at least 2 sleep onset REM periods (REM latency of less than or equal to 15 minutes in at least 2 naps).

| Outcome | Split | Positive Cases (%) | Negative Cases (%) | Median Days To Event (IQR) | Minimum Event Year | Maximum Event Year | Minimum Days to Event | Maximum Days to Event |
|---|---|---|---|---|---|---|---|---|
| Angina | Train | 353 (8.2%) | 3969 (91.8%) | 1645.7 (661.0-2453.0) | 0 | 13 | 31 | 4760 |
| | Val | 48 (10.0%) | 431 (90.0%) | 1307.3 (364.0-2056.5) | 0 | 11 | 21 | 4212 |
| | Test | 96 (8.0%) | 1108 (92.0%) | 1459.5 (505.0-2279.8) | 0 | 11 | 20 | 4103 |
| Cardiovascular Disease Death | Train | 804 (14.8%) | 4616 (85.2%) | 3365.6 (1935.8-4594.5) | 0 | 19 | 7 | 7299 |
| | Val | 88 (14.6%) | 515 (85.4%) | 3497.2 (2323.5-4816.8) | 0 | 19 | 99 | 7127 |
| | Test | 210 (13.9%) | 1296 (86.1%) | 3505.4 (2121.5-4799.8) | 0 | 19 | 42 | 7113 |
| | Ind Test | 116 (5.6%) | 1938 (94.4%) | 2455.1 (1609.0-3324.3) | 0 | 10 | 135 | 3978 |
| Congestive Heart Failure | Train | 582 (10.3%) | 5059 (89.7%) | 1989.3 (1015.0-2963.5) | 0 | 12 | 15 | 4580 |
| | Val | 51 (8.1%) | 577 (91.9%) | 1736.7 (1075.0-2448.0) | 0 | 11 | 9 | 4134 |
| | Test | 166 (10.6%) | 1401 (89.4%) | 1992.4 (972.8-2917.2) | 0 | 12 | 7 | 4608 |
| Coronary Heart Disease Death | Train | 234 (4.2%) | 5381 (95.8%) | 2179.6 (1235.0-3149.0) | 0 | 12 | 85 | 4410 |
| | Val | 35 (5.6%) | 590 (94.4%) | 2187.0 (819.5-3325.0) | 0 | 12 | 159 | 4491 |
| | Test | 68 (4.4%) | 1486 (95.6%) | 2145.6 (1363.0-2884.0) | 0 | 11 | 78 | 4236 |
| | Ind Test | 53 (2.6%) | 2001 (97.4%) | 2449.9 (1369.0-3306.0) | 1 | 10 | 514 | 3978 |
| Incident Cognitive Decline | Train | 386 (18.5%) | 1698 (81.5%) | 1629.7 (476.0-2393.0) | 0 | 11 | 104 | 4373 |
| | Val | 50 (21.6%) | 182 (78.4%) | 1637.2 (486.8-2398.5) | 0 | 11 | 145 | 4359 |
| | Test | 111 (19.1%) | 470 (80.9%) | 1501.4 (468.0-2280.5) | 0 | 12 | 101 | 4459 |
| Incident Diabetes | Train | 167 (4.7%) | 3416 (95.3%) | 1921.1 (1314.5-2327.0) | 0 | 12 | 153 | 4397 |
| | Val | 23 (5.8%) | 376 (94.2%) | 1812.8 (1397.0-1997.5) | 1 | 10 | 392 | 3719 |
| | Test | 52 (5.2%) | 947 (94.8%) | 1860.5 (1239.8-1957.0) | 0 | 10 | 321 | 3938 |
| Incident Hypertension | Train | 626 (19.6%) | 2562 (80.4%) | 1811.4 (1287.2-1977.8) | 1 | 11 | 379 | 4264 |
| | Val | 60 (16.9%) | 296 (83.1%) | 1836.1 (1444.5-1987.5) | 1 | 11 | 528 | 4074 |
| | Test | 174 (19.6%) | 713 (80.4%) | 1927.1 (1393.2-2006.5) | 1 | 11 | 458 | 4264 |
| Incident OSA | Train | 281 (13.0%) | 1879 (87.0%) | 2081.5 (1877.0-2311.0) | 4 | 7 | 1735 | 2812 |
| | Val | 33 (13.8%) | 207 (86.2%) | 2042.5 (1856.0-2118.0) | 4 | 7 | 1756 | 2750 |
| | Test | 81 (13.5%) | 518 (86.5%) | 2047.0 (1868.0-2120.0) | 4 | 7 | 1741 | 2746 |
| | Ind Test | 122 (20.6%) | 471 (79.4%) | 2218.4 (1461.0-2922.0) | 1 | 14 | 365.25 | 5113.5 |
| Myocardial Infarction | Train | 273 (4.8%) | 5365 (95.2%) | 2013.0 (1082.0-2998.0) | 0 | 12 | 21 | 4473 |
| | Val | 36 (5.7%) | 592 (94.3%) | 1825.2 (831.5-2541.8) | 0 | 12 | 229 | 4615 |
| | Test | 68 (4.3%) | 1502 (95.7%) | 2131.1 (1127.5-3026.8) | 0 | 12 | 19 | 4747 |
| | Ind Test | 84 (4.2%) | 1926 (95.8%) | 1903.9 (1081.5-2650.0) | 0 | 11 | 5 | 4104 |
| Stroke | Train | 344 (6.1%) | 5292 (93.9%) | 1801.9 (919.5-2620.2) | 0 | 14 | 4 | 5154 |
| | Val | 29 (4.6%) | 600 (95.4%) | 1481.7 (649.0-2005.0) | 0 | 12 | 109 | 4539 |
| | Test | 90 (5.7%) | 1481 (94.3%) | 1761.4 (696.8-2482.2) | 0 | 12 | 18 | 4572 |
| | Ind Test | 70 (3.5%) | 1949 (96.5%) | 1718.6 (871.3-2517.0) | 0 | 10 | 54 | 3962 |

Supplementary Table 3: Long-term disease risk training (train), validation (val), testing (test), and independent test set (ind test) distributions of events and times to events for finetuning. Outcomes were provided by the study cohort datasets. “Incident” outcomes were calculated manually from follow up visits. Training, validation, and held-out testing followed identical splitting ratios for all outcomes: 72%, 8%, 20%, respectively. Differences in prevalence of events and distributions of time to events are notable between independent test sets and training sets and could affect performance.

| Outcome | Year | IBS<br>SleepJEPA+ | SleepJEPA | Age+Sex |
|---|---|---|---|---|
| Angina | 1 | 0.013 | 0.012 | 0.012 |
| | 2 | 0.021 | 0.019 | 0.019 |
| | 3 | 0.027 | 0.026 | 0.025 |
| | 5 | 0.035 | 0.035 | 0.034 |
| | 10 | 0.05 | 0.053 | 0.051 |
| | 15 | 0.058 | 0.064 | 0.058 |
| CV Disease Death | 1 | 0.003 | 0.006 | 0.003 |
| | 2 | 0.006 | 0.009 | 0.006 |
| | 3 | 0.008 | 0.011 | 0.008 |
| | 5 | 0.013 | 0.016 | 0.013 |
| | 10 | 0.031 | 0.034 | 0.031 |
| | 15 | 0.055 | 0.062 | 0.053 |
| Congestive Heart Failure | 1 | 0.011 | 0.011 | 0.011 |
| | 2 | 0.016 | 0.016 | 0.016 |
| | 3 | 0.02 | 0.02 | 0.021 |
| | 5 | 0.028 | 0.028 | 0.029 |
| | 10 | 0.047 | 0.048 | 0.05 |
| | 15 | 0.06 | 0.066 | 0.065 |
| Coronary Heart Disease Death | 1 | 0.002 | 0.002 | 0.002 |
| | 2 | 0.004 | 0.004 | 0.004 |
| | 3 | 0.006 | 0.006 | 0.006 |
| | 5 | 0.009 | 0.009 | 0.009 |
| | 10 | 0.021 | 0.021 | 0.021 |
| | 15 | 0.03 | 0.031 | 0.029 |
| Incident Cognitive Decline | 1 | 0.014 | 0.014 | 0.014 |
| | 2 | 0.045 | 0.045 | 0.044 |
| | 3 | 0.063 | 0.061 | 0.06 |
| | 5 | 0.077 | 0.073 | 0.073 |
| | 10 | 0.09 | 0.086 | 0.085 |
| Incident Diabetes | 1 | 0.001 | 0.001 | 0.001 |
| | 2 | 0.005 | 0.005 | 0.005 |
| | 3 | 0.007 | 0.007 | 0.007 |
| | 5 | 0.012 | 0.012 | 0.011 |
| | 10 | 0.037 | 0.038 | 0.037 |
| Incident Hypertension | 2 | 0.016 | 0.016 | 0.016 |
| | 3 | 0.025 | 0.026 | 0.025 |
| | 5 | 0.05 | 0.052 | 0.048 |
| | 10 | 0.106 | 0.107 | 0.127 |
| Incident OSA | 5 | 0.016 | 0.016 | 0.022 |
| Myocardial Infarction | 1 | 0.002 | 0.002 | 0.002 |
| | 2 | 0.004 | 0.004 | 0.004 |
| | 3 | 0.006 | 0.006 | 0.006 |
| | 5 | 0.01 | 0.01 | 0.01 |
| | 10 | 0.022 | 0.021 | 0.021 |
| | 15 | 0.033 | 0.032 | 0.031 |
| Stroke | 1 | 0.004 | 0.004 | 0.004 |
| | 2 | 0.01 | 0.01 | 0.009 |
| | 3 | 0.013 | 0.013 | 0.013 |
| | 5 | 0.02 | 0.02 | 0.02 |
| | 10 | 0.034 | 0.034 | 0.033 |
| | 15 | 0.045 | 0.044 | 0.041 |

Supplementary Table 4: SleepJEPA long-term outcomes integrated Brier score (IBS) at different forecast lengths on the 20% held-out test sets. SleepJEPA+, SleepJEPA, and the baseline demographics models are reported.

| Outcome | Dataset | Year | IBS<br>SleepJEPA+ | SleepJEPA | Age+Sex |
|---|---|---|---|---|---|
| CV Disease Death | MESA | 1 | 0.001 | 0.005 | 0.001 |
| | | 2 | 0.002 | 0.007 | 0.002 |
| | | 3 | 0.005 | 0.01 | 0.005 |
| | | 5 | 0.009 | 0.014 | 0.009 |
| | | 10 | 0.022 | 0.027 | 0.021 |
| Coronary Heart Disease Death | MESA | 2 | 0.002 | 0.002 | 0.002 |
| | | 3 | 0.003 | 0.003 | 0.003 |
| | | 5 | 0.005 | 0.006 | 0.005 |
| | | 10 | 0.012 | 0.012 | 0.011 |
| Incident OSA | WSC | 5 | 0.046 | 0.045 | 0.042 |
| | | 10 | 0.368 | 0.088 | 0.143 |
| | | 15 | 0.237 | 0.057 | 0.093 |
| Myocardial Infarction | MESA | 1 | 0.002 | 0.002 | 0.002 |
| | | 2 | 0.005 | 0.005 | 0.005 |
| | | 3 | 0.007 | 0.007 | 0.007 |
| | | 5 | 0.012 | 0.012 | 0.012 |
| | | 10 | 0.024 | 0.024 | 0.023 |
| Stroke | MESA | 1 | 0.004 | 0.004 | 0.004 |
| | | 2 | 0.006 | 0.006 | 0.006 |
| | | 3 | 0.007 | 0.007 | 0.007 |
| | | 5 | 0.011 | 0.011 | 0.011 |
| | | 10 | 0.021 | 0.021 | 0.021 |

Supplementary Table 5: SleepJEPA long-term outcomes integrated Brier score at different forecast lengths on independent test sets, if available. SleepJEPA+, SleepJEPA, and the baseline demographics models are reported.

| Outcome | Variable | Average Correlation (95% CI) |
|---|---|---|
| Angina | Age | 0.38 (0.37 - 0.4) |
| | Central AHI | 0.25 (0.23 - 0.26) |
| | Obstructive AHI | 0.23 (0.21 - 0.24) |
| | Total sleep time | -0.2 (-0.22 - -0.19) |
| | Avg SpO2 | -0.19 (-0.2 - -0.17) |
| | Sleep efficiency | -0.17 (-0.18 - -0.16) |
| | REM sleep time | -0.15 (-0.16 - -0.14) |
| | Arousal index | 0.15 (0.14 - 0.16) |
| | Min SpO2 | -0.14 (-0.16 - -0.13) |
| | N1 sleep time | 0.08 (0.08 - 0.09) |
| | N2 sleep time | 0.08 (0.08 - 0.09) |
| | BMI | 0.08 (0.07 - 0.09) |
| | N3 sleep time | -0.03 (-0.04 - -0.03) |
| | ESS | 0.01 (0.01 - 0.02) |
| Cardiovascular Disease Death | Min SpO2 | -0.41 (-0.42 - -0.41) |
| | Obstructive AHI | 0.39 (0.38 - 0.39) |
| | Age | 0.36 (0.35 - 0.37) |
| | Central AHI | 0.35 (0.34 - 0.36) |
| | Avg SpO2 | -0.31 (-0.32 - -0.31) |
| | Sleep efficiency | -0.3 (-0.31 - -0.29) |
| | Total sleep time | -0.25 (-0.25 - -0.24) |
| | N1 sleep time | 0.23 (0.22 - 0.23) |
| | Arousal index | 0.2 (0.19 - 0.21) |
| | REM sleep time | -0.15 (-0.16 - -0.15) |
| | N2 sleep time | 0.07 (0.06 - 0.07) |
| | BMI | 0.06 (0.06 - 0.07) |
| | N3 sleep time | -0.06 (-0.07 - -0.05) |
| | ESS | -0.02 (-0.02 - -0.01) |
| Congestive Heart Failure | Age | 0.39 (0.37 - 0.4) |
| | Avg SpO2 | -0.34 (-0.35 - -0.33) |
| | Central AHI | 0.28 (0.27 - 0.29) |
| | Min SpO2 | -0.26 (-0.28 - -0.25) |
| | Sleep efficiency | -0.25 (-0.26 - -0.25) |
| | Obstructive AHI | 0.24 (0.23 - 0.26) |
| | Arousal index | 0.2 (0.19 - 0.21) |
| | Total sleep time | -0.18 (-0.18 - -0.18) |
| | N1 sleep time | 0.15 (0.15 - 0.15) |
| | REM sleep time | -0.13 (-0.13 - -0.13) |
| | N2 sleep time | 0.1 (0.1 - 0.11) |
| | N3 sleep time | -0.09 (-0.09 - -0.08) |
| | BMI | 0.05 (0.04 - 0.06) |
| | ESS | -0.01 (-0.01 - 0) |
| Coronary Heart Disease Death | Obstructive AHI | 0.3 (0.28 - 0.31) |
| | Avg SpO2 | -0.27 (-0.28 - -0.26) |
| | Arousal index | 0.27 (0.24 - 0.3) |
| | Min SpO2 | -0.25 (-0.27 - -0.24) |
| | Age | 0.25 (0.24 - 0.26) |
| | Sleep efficiency | -0.19 (-0.2 - -0.18) |
| | Central AHI | 0.17 (0.16 - 0.17) |
| | REM sleep time | -0.12 (-0.13 - -0.11) |
| | N1 sleep time | 0.12 (0.11 - 0.12) |
| | N2 sleep time | 0.11 (0.1 - 0.12) |
| | Total sleep time | -0.11 (-0.12 - -0.1) |
| | N3 sleep time | -0.09 (-0.09 - -0.08) |
| | ESS | 0.05 (0.04 - 0.05) |

| | | |
|---|---|---|
| | BMI | 0.04 (0.03 - 0.04) |
| Incident Cognitive Decline | Age | 0.21 (0.2 - 0.21) |
| | Sleep efficiency | -0.2 (-0.22 - -0.19) |
| | REM sleep time | -0.16 (-0.18 - -0.15) |
| | Total sleep time | -0.14 (-0.16 - -0.13) |
| | Central AHI | -0.08 (-0.09 - -0.07) |
| | Arousal index | 0.07 (0.06 - 0.09) |
| | N2 sleep time | -0.07 (-0.08 - -0.06) |
| | BMI | -0.06 (-0.07 - -0.04) |
| | ESS | 0.04 (0.03 - 0.04) |
| | Min SpO2 | 0.04 (0.02 - 0.06) |
| | N1 sleep time | -0.03 (-0.04 - -0.02) |
| | Obstructive AHI | 0.03 (0 - 0.05) |
| | Avg SpO2 | 0.02 (0 - 0.04) |
| | N3 sleep time | -0.02 (-0.03 - 0) |
| Incident Diabetes | Avg SpO2 | -0.32 (-0.38 - -0.27) |
| | Obstructive AHI | 0.32 (0.29 - 0.35) |
| | Central AHI | 0.31 (0.28 - 0.33) |
| | Min SpO2 | -0.23 (-0.25 - -0.22) |
| | BMI | 0.22 (0.18 - 0.25) |
| | Arousal index | 0.21 (0.18 - 0.25) |
| | Sleep efficiency | -0.18 (-0.2 - -0.15) |
| | N1 sleep time | 0.14 (0.11 - 0.17) |
| | Total sleep time | -0.12 (-0.14 - -0.1) |
| | Age | 0.11 (0.1 - 0.12) |
| | N3 sleep time | -0.1 (-0.12 - -0.07) |
| | N2 sleep time | 0.09 (0.06 - 0.11) |
| | REM sleep time | -0.05 (-0.07 - -0.04) |
| | ESS | 0.01 (-0.02 - 0.04) |
| Incident Hypertension | Age | 0.24 (0.21 - 0.27) |
| | BMI | 0.18 (0.15 - 0.2) |
| | Min SpO2 | -0.16 (-0.16 - -0.15) |
| | Central AHI | 0.15 (0.14 - 0.15) |
| | Obstructive AHI | 0.14 (0.13 - 0.14) |
| | Avg SpO2 | -0.13 (-0.14 - -0.12) |
| | Total sleep time | -0.13 (-0.13 - -0.12) |
| | Sleep efficiency | -0.12 (-0.13 - -0.12) |
| | REM sleep time | -0.05 (-0.06 - -0.03) |
| | N1 sleep time | 0.04 (0.03 - 0.05) |
| | ESS | -0.04 (-0.07 - 0) |
| | Arousal index | 0.03 (0.02 - 0.04) |
| | N3 sleep time | 0.01 (-0.01 - 0.02) |
| | N2 sleep time | 0.01 (0 - 0.01) |
| Incident OSA | Avg SpO2 | -0.41 (-0.46 - -0.36) |
| | Obstructive AHI | 0.41 (0.36 - 0.47) |
| | Central AHI | 0.39 (0.34 - 0.44) |
| | Min SpO2 | -0.32 (-0.37 - -0.28) |
| | BMI | 0.22 (0.2 - 0.24) |
| | Age | 0.18 (0.15 - 0.21) |
| | Total sleep time | -0.16 (-0.17 - -0.14) |
| | Arousal index | 0.16 (0.13 - 0.18) |
| | REM sleep time | -0.12 (-0.14 - -0.11) |
| | Sleep efficiency | -0.1 (-0.12 - -0.07) |
| | N2 sleep time | 0.06 (0.05 - 0.07) |
| | N1 sleep time | 0.06 (0.05 - 0.07) |
| | ESS | 0.06 (0.05 - 0.07) |

| | | |
|---|---|---|
| | N3 sleep time | -0.02 (-0.03 - 0) |
| Myocardial Infarction | Obstructive AHI | 0.29 (0.27 - 0.31) |
| | Avg SpO2 | -0.27 (-0.29 - -0.24) |
| | Central AHI | 0.26 (0.24 - 0.28) |
| | Age | 0.23 (0.21 - 0.24) |
| | Min SpO2 | -0.21 (-0.23 - -0.19) |
| | N2 sleep time | 0.14 (0.13 - 0.15) |
| | Arousal index | 0.14 (0.12 - 0.15) |
| | N3 sleep time | -0.14 (-0.14 - -0.13) |
| | Sleep efficiency | -0.12 (-0.13 - -0.1) |
| | N1 sleep time | 0.08 (0.08 - 0.08) |
| | BMI | 0.08 (0.07 - 0.08) |
| | ESS | 0.06 (0.06 - 0.07) |
| | REM sleep time | -0.05 (-0.07 - -0.04) |
| | Total sleep time | 0 (-0.01 - 0.01) |
| Stroke | Age | 0.37 (0.35 - 0.38) |
| | Sleep efficiency | -0.21 (-0.22 - -0.2) |
| | Arousal index | 0.19 (0.18 - 0.2) |
| | Total sleep time | -0.18 (-0.19 - -0.17) |
| | Central AHI | 0.18 (0.16 - 0.19) |
| | Obstructive AHI | 0.17 (0.16 - 0.18) |
| | Avg SpO2 | -0.17 (-0.19 - -0.15) |
| | REM sleep time | -0.15 (-0.16 - -0.14) |
| | Min SpO2 | -0.14 (-0.15 - -0.14) |
| | N1 sleep time | 0.06 (0.06 - 0.07) |
| | BMI | 0.04 (0.03 - 0.04) |
| | N2 sleep time | 0.04 (0.03 - 0.04) |
| | N3 sleep time | 0.02 (0.02 - 0.03) |
| | ESS | 0.01 (0 - 0.01) |

Supplementary Table 6: SleepJEPA cumulative risk predictions average Pearson correlations across all risk horizons from the SHHS or MrOS (for cognitive decline) held-out test set. Each outcome is sorted from highest to lowest absolute correlation. 95% confidence intervals are reported.

#### References


M Brandon Westover, Valdery Moura Junior, Robert Thomas, Sydney Cash, Samaneh Nasiri, Haoqi Sun, Aditya Gupta, Jonathan Rosand, Manohar Ghanta, Wolfgang Ganglberger, Umakanth Katwa, Katie Stone, Zhiyong Zhang, Gauri Ganjoo, Thijs E Nassi PhD Candidate, Ruoqi Wei, Dennis Hwang, Lynn Marie Trotti, Ankit Parekh, ErikJan Meulenbrugge, Emmanuel Mignot, Rhoda Au, Gari Clifford, and David Rapoport. The Human Sleep Project. URL `https://bdsp.io/content/hsp/2.0/`.

S. F. Quan, B. V. Howard, C. Iber, J. P. Kiley, F. J. Nieto, G. T. O'Connor, D. M. Rapoport, S. Redline, J. Robbins, J. M. Samet, and P. W. Wahl. The Sleep Heart Health Study: design, rationale, and methods. *Sleep*, 20(12):1077–1085, December 1997. ISSN 0161-8105.

Guo-Qiang Zhang, Licong Cui, Remo Mueller, Shiqiang Tao, Matthew Kim, Michael Rueschman, Sara Mariani, Daniel Mobley, and Susan Redline. The National Sleep Research Resource: towards a sleep data commons. *Journal of the American Medical Informatics Association: JAMIA*, 25(10):1351–1358, October 2018. ISSN 1527-974X. doi:10.1093/jamia/ocy064.

Terry Young. Rationale, design and findings from the Wisconsin Sleep Cohort Study: Toward understanding the total societal burden of sleep disordered breathing. *Sleep medicine clinics*, 4(1):37–46, March 2009. ISSN 1556-407X. doi:10.1016/j.jsmc.2008.11.003. URL `https://www.ncbi.nlm.nih.gov/pmc/articles/PMC2699290/`.

Terri Blackwell, Kristine Yaffe, Sonia Ancoli-Israel, Susan Redline, Kristine E. Ensrud, Marcia L. Stefanick, Alison Laffan, Katie L. Stone, and Osteoporotic Fractures in Men (MrOS) Study Group. Association of sleep characteristics and cognition in older community-dwelling men: the MrOS sleep study. *Sleep*, 34(10):1347–1356, October 2011. ISSN 1550-9109. doi:10.5665/SLEEP.1276.

Xiaoli Chen, Rui Wang, Phyllis Zee, Pamela L. Lutsey, Sogol Javaheri, Carmela Alcántara, Chandra L. Jackson, Michelle A. Williams, and Susan Redline. Racial/Ethnic Differences in Sleep Disturbances: The Multi-Ethnic Study of Atherosclerosis (MESA). *Sleep*, 38(6):877–888, June 2015. ISSN 1550-9109. doi:10.5665/sleep.4732.

Jens B. Stephansen, Alexander N. Olesen, Mads Olsen, Aditya Ambati, Eileen B. Leary, Hyatt E. Moore, Oscar Carrillo, Ling Lin, Fang Han, Han Yan, Yun L. Sun, Yves Dauvilliers, Sabine Scholz, Lucie Barateau, Birgit Hogl, Ambra Stefani, Seung Chul Hong, Tae Won Kim, Fabio Pizza, Giuseppe Plazzi, Stefano Vandi, Elena Antelmi, Dimitri Perrin, Samuel T. Kuna, Paula K. Schweitzer, Clete Kushida, Paul E. Peppard, Helge B. D. Sorensen, Poul Jennum, and Emmanuel Mignot. Neural network analysis of sleep stages enables efficient diagnosis of narcolepsy. *Nature Communications*, 9(1):5229, December 2018. ISSN 2041-1723. doi:10.1038/s41467-018-07229-3. URL `https://www.nature.com/articles/s41467-018-07229-3`.

Stuart F. Quan, Cynthia S. Chan, William C. Dement, Alan Gevins, James L. Goodwin, Daniel J. Gottlieb, Sylvan Green, Christian Guilleminault, Max Hirshkowitz, Pamela R. Hyde, Gary G. Kay, Eileen B. Leary, Deborah A. Nichols, Paula K. Schweitzer, Richard D. Simon, James K. Walsh, and Clete A. Kushida. The Association between Obstructive Sleep Apnea and Neurocognitive Performance—The Apnea Positive Pressure Long-term Efficacy Study (APPLES). *Sleep*, 34(3):303. doi:10.1093/sleep/34.3.303. URL `https://www.ncbi.nlm.nih.gov/pmc/articles/PMC3041706/`.

Alistair Miles, jakirkham, M. Bussonnier, Josh Moore, Dimitri Papadopoulos Orfanos, Davis Bennett, David Stansby, Joe Hamman, James Bourbeau, Andrew Fulton, Gregory Lee, Ryan Abernathey, Norman Rzepka, Zain Patel, Mads R. B. Kristensen, Sanket Verma, Saransh Chopra, Matthew Rocklin, AWA BRANDON AWA, Max Jones, Martin Durant, Elliott Sales de Andrade, Vincent Schut, raphael dussin, Shivank Chaudhary, Chris Barnes, Juan Nunez-Iglesias, and shikharsg. zarr-developers/zarr-python: v3.0.0-alpha, June 2024. URL `https://zenodo.org/records/11592827`.

Andreas Brink-Kjaer, Eileen B. Leary, Haoqi Sun, M. Brandon Westover, Katie L. Stone, Paul E. Peppard, Nancy E. Lane, Peggy M. Cawthon, Susan Redline, Poul Jennum, Helge B. D. Sorensen, and Emmanuel Mignot. Age estimation from sleep studies using deep learning predicts life expectancy. *npj Digital Medicine*, 5(1):1–10, July 2022. ISSN 2398-6352. doi:10.1038/s41746-022-00630-9. URL `https://www.nature.com/articles/s41746-022-00630-9`.

Adam Paszke, Sam Gross, Francisco Massa, Adam Lerer, James Bradbury, Gregory Chanan, Trevor Killeen, Zeming Lin, Natalia Gimelshein, Luca Antiga, Alban Desmaison, Andreas Köpf, Edward Yang, Zach DeVito, Martin Raison, Alykhan Tejani, Sasank Chilamkurthy, Benoit Steiner, Lu Fang, Junjie Bai, and Soumith Chintala. PyTorch: an imperative style, high-performance deep learning library. In *Proceedings of the 33rd International Conference on Neural Information Processing Systems*, number 721, pages 8026–8037. Curran Associates Inc., Red Hook, NY, USA, December 2019. URL `https://dl.acm.org/doi/10.5555/3454287.3455008`.

Bastien Lechat, Kristy L. Hansen, Yohannes Adama Melaku, Andrew Vakulin, Gorica Micic, Robert J. Adams, Sarah Appleton, Danny J. Eckert, Peter Catcheside, and Branko Zajamsek. A Novel Electroencephalogram-derived

Measure of Disrupted Delta Wave Activity during Sleep Predicts All-Cause Mortality Risk. *Annals of the American Thoracic Society*, 19(4):649–658, April 2022. ISSN 2329-6933. doi:10.1513/AnnalsATS.202103-315OC. URL https://www.atsjournals.org/doi/10.1513/AnnalsATS.202103-315OC.

Magdy Younes, Michele Ostrowski, Marc Soiferman, Henry Younes, Mark Younes, Jill Raneri, and Patrick Hanly. Odds Ratio Product of Sleep EEG as a Continuous Measure of Sleep State. *Sleep*, 38(4):641–654, April 2015. ISSN 0161-8105. doi:10.5665/sleep.4588. URL https://doi.org/10.5665/sleep.4588.

Mingze Dong, Leda Wang, and Yuval Kluger. Understanding and Enhancing Mask-Based Pretraining towards Universal Representations, September 2025. URL http://arxiv.org/abs/2509.21650. arXiv:2509.21650 [cs].

Benjamin Fox, Dung T. Hoang, Joy Jiang, Pushkala Jayaraman, Ankit Parekh, Girish N. Nadkarni, and Ankit Sakhuja. PhysioJEPA: Joint Embedding Representations of Physiological Signals for Real Time Risk Estimation in the Intensive Care Unit. November 2025. URL https://openreview.net/forum?id=bdXsfrNaGY.

Yuqi Nie, Nam H. Nguyen, Phanwadee Sinthong, and Jayant Kalagnanam. A Time Series is Worth 64 Words: Long-term Forecasting with Transformers. In *The Eleventh International Conference on Learning Representations*, 2023. URL https://openreview.net/forum?id=Jbdc0vTOcol.

Jianlin Su, Murtadha Ahmed, Yu Lu, Shengfeng Pan, Wen Bo, and Yunfeng Liu. RoFormer: Enhanced transformer with Rotary Position Embedding. *Neurocomput.*, 568(C), February 2024. ISSN 0925-2312. doi:10.1016/j.neucom.2023.127063. URL https://doi.org/10.1016/j.neucom.2023.127063.

Edward A. Wolpert. A Manual of Standardized Terminology, Techniques and Scoring System for Sleep Stages of Human Subjects. *Archives of General Psychiatry*, 20(2):246–247, February 1969. ISSN 0003-990X. doi:10.1001/archpsyc.1969.01740140118016. URL https://doi.org/10.1001/archpsyc.1969.01740140118016.

E. L. Teng and H. C. Chui. The Modified Mini-Mental State (3MS) examination. *The Journal of Clinical Psychiatry*, 48(8):314–318, August 1987. ISSN 0160-6689.

Clémence Cavaillès, Meredith Wallace, Yue Leng, Katie L. Stone, Sonia Ancoli-Israel, and Kristine Yaffe. Multidimensional sleep profiles via machine learning and risk of dementia and cardiovascular disease. *Communications Medicine*, 5(1):306, July 2025. ISSN 2730-664X. doi:10.1038/s43856-025-01019-x. URL https://www.nature.com/articles/s43856-025-01019-x.
